# Enhancement of Sleep Slow Wave Activity using Transcranial Electrical Stimulation with Temporal Interference

**DOI:** 10.1101/2025.08.11.25333452

**Authors:** Erin L. Schaeffer, Ido Haber, Zhiwei Fan, Simone Bruno, Beril Mat, Tariq Alauddin, Giulietta Vigueras, Lindsey Neumann, Richard Smith, Florian Missey, Adam Williamson, Peter Achermann, Stefan Beerli, Myles Capstick, Esra Neufeld, Niels Kuster, Robin I. Goldman, Richard J. Davidson, Larissa Albantakis, Stephanie G. Jones, Chiara Cirelli, Melanie Boly, Giulio Tononi

**Affiliations:** Department of Psychiatry, University of Wisconsin-Madison, Madison, Wisconsin; Medical Scientist Training Program, University of Wisconsin-Madison, Madison, Wisconsin; Department of Biomedical Engineering, University of Wisconsin-Madison, Madison, Wisconsin; International Institute for Integrative Sleep Medicine (WPI-IIIS), Tsukuba Institute for Advanced Research (TIAR), University of Tsukuba, Tsukuba, Japan; International Clinical Research Center, St. Anne’s University Hospital Brno, Brno, Czech Republic; Institute de Neurosciences des Systèmes (INS), INSERM, Aix-Marseille Université, Marseille, France; Institute of Pharmacology and Toxicology, University of Zurich, Zurich, Switzerland; Foundation for Research on Information Technologies in Society (IT’IS), Zurich, Switzerland; Department of Information Technology and Electrical Engineering, Swiss Federal Institute of Technology (ETH) Zurich, Zurich, Switzerland; Center for Healthy Minds, University of Wisconsin-Madison, Madison, Wisconsin; Department of Psychology, University of Wisconsin-Madison, Madison, Wisconsin; Department of Neurology, University of Wisconsin-Madison, Madison, Wisconsin

## Abstract

Slow waves mediate restorative functions of non-rapid eye movement (NREM) sleep. To enhance slow wave activity (SWA, 0.5-4 Hz) non-invasively, we employed a novel neuromodulatory tool, Transcranial Electrical Stimulation with Temporal Interference (TES-TI). Healthy participants (n=21) received TES-TI during NREM sleep over several nights (∼10 stimulation periods, 3min each, first half of night, 4-week protocol), targeting left ventromedial prefrontal cortex—a “hot spot” of slow wave generation. Two high frequency carriers with 1Hz difference (TES^15kHz^-TI^1Hz^) produced amplitude modulated temporal interference at 1Hz. SWA, measured with simultaneous high-density electroencephalography (hd-EEG), was increased by TES^15kHz^-TI^1Hz^, with the effect outlasting the stimulation period. Higher frequencies (sigma and beta) decreased. Pure TES^15kHz^ did not have comparable effects. The incremental effects of TES^15kHz^-TI^1Hz^ on SWA between first and last intervention night were positively correlated with subjective ratings of restorative sleep. This is the first study demonstrating that TES-TI can enhance sleep and its restorative function.

## Introduction

During non-rapid eye movement (NREM) sleep, which comprises 75–80% of total time spent in sleep, individual slow waves are detected on the EEG when many cortical neurons switch near-synchronously between ON periods of firing and OFF periods of silence ^1^. Sleep slow waves are homeostatically regulated. Thus, slow wave activity (SWA, electroencephalographic power in 0.5–4 Hz range), a combined measure of the number and amplitude of slow waves, is high in early sleep, even higher after sleep deprivation, and declines in the course of sleep when sleep need is discharged ^2-4^. Slow waves are believed to mediate key restorative functions attributed to NREM sleep, including synaptic renormalization and memory consolidation ^5-8^. Disrupted SWA is associated with worsened cognitive functioning, including poorer performance on visuomotor and perceptual learning tasks ^9-11^. Conversely, the enhancement of SWA has been shown to improve memory performance ^12-16^.

Two interventions commonly used to enhance SWA are acoustic stimulation and transcranial electrical stimulation (TES) ^12-14^. However, both techniques have limitations. Acoustic stimulation strong enough to elicit SWA can be close to the threshold for awakening the participant ^10^. Conventional TES, which typically employs frequencies below 1 kHz, results in electrical artifacts during stimulation that prevent adequate evaluation of simultaneous EEG recordings, particularly at the stimulation frequency ^14^. In addition, conventional TES stimulates broadly, affecting superficial regions with diminishing efficacy over deeper brain regions when using parameters that do not produce scalp discomfort ^17^.

An innovative neuromodulatory tool potentially able to overcome these limitations is Transcranial Electrical Stimulation with Temporal Interference (TES-TI) ^18^. It uses two or more high frequency alternating current carriers (e.g., 15 kHz and 15 kHz + 1 Hz) to produce “temporal interference,” that is, amplitude modulation at the difference frequency (e.g., 1 Hz) capable of modulating neural activity at a chosen location in an individual’s brain ^18^. This approach achieves both depth of penetration and focality ^18^. Moreover, TES-TI is steerable, in a way not easily attainable with other non-invasive neuromodulation techniques, by adjusting the current ratio of the two channel pairs ^18^. Importantly, high frequency carriers allow for effective stimulation intensities without the participant perceiving any peripheral stimulation that could cause awakening from sleep ^19,20^. Several studies have now implemented TES-TI with high frequency carriers (up to 20 kHz) in a safe and effective manner in humans−for example, in studies of cognitive tasks ^21-26^−but none of them have used this method to promote sleep and, more specifically, to enhance SWA.

The current study presents data from simultaneously recorded hd-EEG and TES-TI during overnight sleep in a laboratory setting in healthy humans. TES^15kHz^-TI^1Hz^ (15 kHz carrier, 1 Hz difference) targeted left ventromedial prefrontal cortical regions given evidence from source modeling ^27^ and invasive recordings ^28^ suggesting that these areas are a “hot spot” for slow wave generation. With hardware filters, simultaneous hd-EEG recording was obtained during TES^15kHz^-TI^1Hz^ delivered during periods of NREM sleep in the first half of the night. The results show that TES^15kHz^-TI^1Hz^ can enhance sleep SWA.

## Results

Twenty-eight participants (30.5±9.9 years of age, 61% Female) from the first five cohorts of the STRENGTHEN clinical trial were included in the current study. As described in the Methods, participants were assigned to one of four groups, receiving interventions over the course of four weeks. Twenty-one participants (Groups 2,3,4) received TES^15kHz^-TI^1Hz^ and seven other participants (Group 1) received TES^15kHz^. All participants had a baseline overnight with polysomnography (PSG) and hd-EEG recordings. Afterwards, overnight interventions consisted of PSG, hd-EEG, and stimulation. Hardware filters allowed for simultaneous hd-EEG recordings and stimulation (**Figure 1A**). TES^15kHz^-TI^1Hz^ protocols performed during NREM sleep had the following parameters: 15,000 Hz carrier frequency, 1 Hz difference frequency, sinusoidal waveform, 5 mA peak to peak, 3-minute duration with 15-second ramping up/down periods **(Figure lB,D).** Electric field modeling was used to target the left ventromedial prefrontal cortical region. The montage chosen (channel 1: E34-E20, channel 2: E70-E95) generated a theoretical electric field that exceeded 0.7 V/m in the target region when simulated for a representative participant **(Figure lC).** Across all TES^15^_kHz_nrnz_ participants (N=21), maximal intensity values in left ventromedial prefrontal cortex grey matter were 0.82±0.09 V/m, ranging 0.62-0.99 V/m. Average intensity values were 0.46±0.05 V/m, ranging 0.36-0.52 V/m. Stimulations occurred in three-minute protocols repeated ten times during NREM sleep identified online by sleep technicians **(Figure 1D).** In most participants, stimulations were completed within the first four hours from sleep onset **(Figure 1D).** Stimulation parameters were the same across groups except that TES^15^_kHz_ had a difference of 0 Hz.

**Figure 1.**
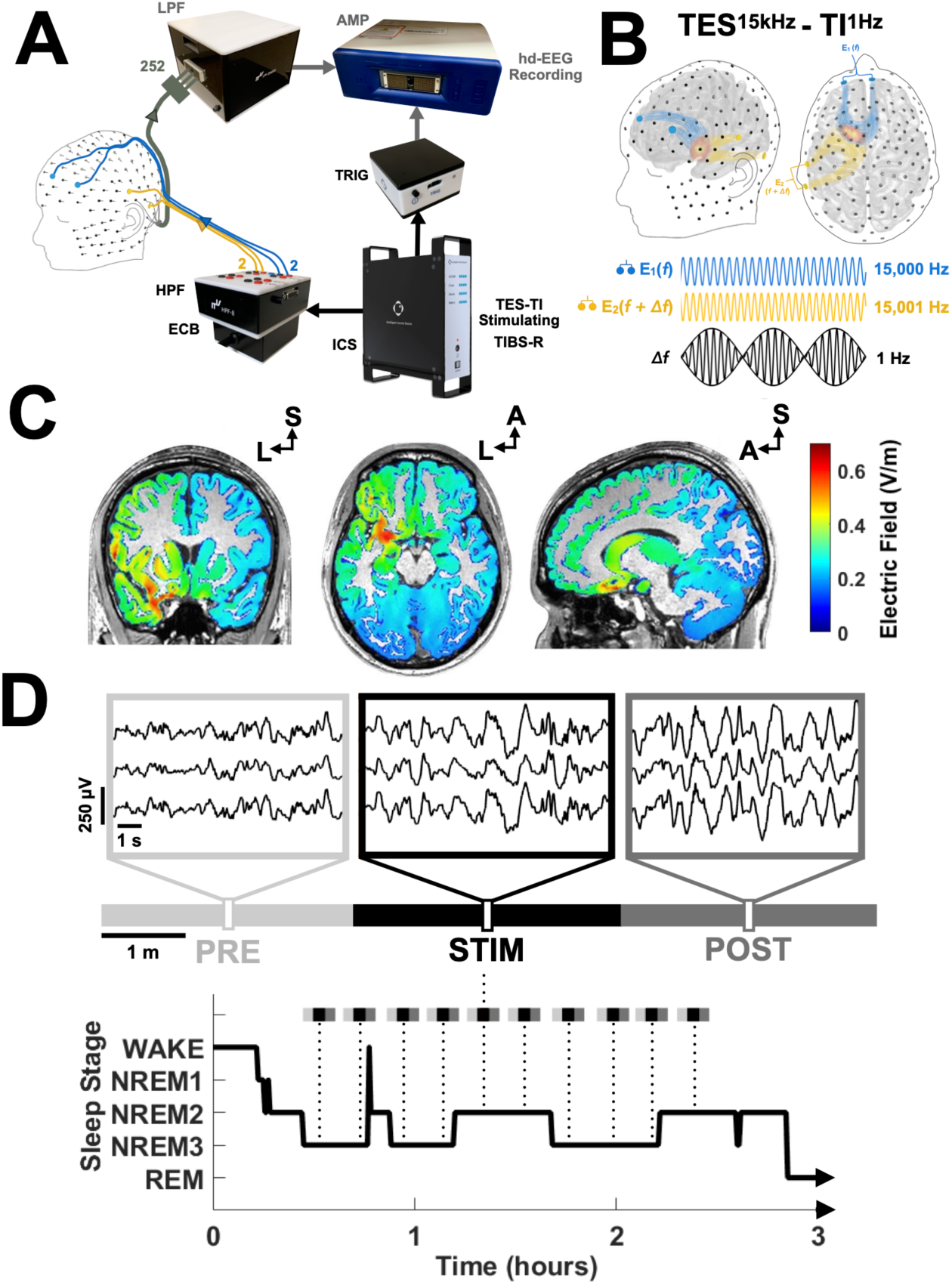
Experiment overview. **A.** The Transcranial Electrical Stimulation with Temporal Interference (TES­ T!) system comprised the Temporal Interference Brain Stimulator for Research (TIBS-R) with components including the Intelligent Current Source (JCS), electrode connector box (ECB), and high-pass filter (HPF). Current was delivered through two pairs of stimulating electrodes (four electrodes total) embedded in the hd-EEG net. All other hd-EEG recording channels (252) passed through a low-pass filter (LPF) before reaching the recording amplifier. The dual HPF and LPF system allowed for simultaneous TES-TI and hd-EEG recording. Trigger (TRIG) outputs from the TIBS-R allowed for identification of when stimulation occurred. **B**. TES^15kHz^-TI^1Hz^ consisted of one pair of electrodes delivering high frequency alternating current (E_1_: 15,000 Hz) and the other pair delivering high frequency alternating current with an offset (E_2_: 15,0001 Hz) to create amplitude modulation at the difference frequency (1 Hz) deep in the brain. **C**. Simulated temporal interference electric field in grey matter for an individual participant. Maximum electric field values occurred in deep, left ventromedial prefrontal cortical regions with values exceeding 0.7 V/m. **D**. (Top) A single stimulation protocol with three-minute periods before (PRE), during (STIM), and after (POST) stimulation. Three channels of example EEG data during ten seconds within each three-minute period shown. (Bottom) Representative example of the set of stimulations during NREM sleep, with ten stimulations occurring within the first hours after sleep onset.

Baseline, first, and last intervention nights were included in the current study. In total, twenty-two first nights (17 TES^15kHz^-TI^1Hz^, 5 TES^15kHz^) and twenty-seven last nights (20 TES^15kHz^-TI^1Hz^, 7 TES^15kHz^) were used in analyses that evaluated effects during stimulation (see Methods for details). One additional first night and one additional last night for a TES^15kHz^-TI^1Hz^ participant were included in analyses that evaluated the effects before or after stimulation. Analyses comparing baseline, first, and last intervention nights were restricted to only participants with usable data on all nights (16 TES^15kHz^-TI^1Hz^, 5 TES^15kHz^).

Across all participants, the age range was 19–49 years, race was 85.7% White, 14.3% Asian, and ethnicity was 96.4% non-Hispanic, 3.6% Hispanic, with no statistically significant differences in age or sex between TES^15kHz^-TI^1Hz^ and TES^15kHz^ (**Table 1**). Sleep characteristics during the baseline night also did not differ between TES^15kHz^-TI^1Hz^ and TES^15kHz^, indicating adequate allocation of participants between groups (**Table 2**).

**Table 1.** Basic demographics. TES^15kHz^-TI^1Hz^ and TES^15kHz^ participants showed no statistically significant differences in age (Wilcoxon rank sum test) or sex (chi-squared test). Mean ± standard deviation.

| | $\text{TES}^{15\text{kHz}}\text{-TI}^{1\text{Hz}}$ | $\text{TES}^{15\text{kHz}}$ | Test Statistic, p-value |
| --- | --- | --- | --- |
| **Number of Participants** | 21 | 7 |  |
| **Age (yrs)** | 31.5 ± 10.0 | 27.4 ± 9.8 | (W) 319.5, 0.441 |
| **Sex (%Female)** | 71.4 | 57.1 | ($\chi^2$) 0.45, 0.503 |

**Table 2.** Baseline sleep characteristics. TES^15kHz^-TI^1Hz^ (N=21) and TES^15kHz^ (N=7) participants showed no statistically significant differences in any sleep characteristics during the baseline night (Wilcoxon rank sum test). Mean ± standard deviation.

| | $TES^{15kHz} - TI^{1Hz}$ | $TES^{15kHz}$ | Test Statistic (W), p-value |
| --- | --- | --- | --- |
| Total Sleep-Wake Period (hr) | 8.18 ± 0.94 | 7.62 ± 1.10 | 328.0, 0.222 |
| Total Sleep Time (hr) | 6.83 ± 0.87 | 6.52 ± 1.25 | 311.5, 0.730 |
| Sleep Onset Latency (min) | 23.10 ± 11.92 | 25.93 ± 14.00 | 293.5, 0.577 |
| Sleep Efficiency (%) | 83.81 ± 8.58 | 85.49 ± 10.85 | 286.0, 0.340 |
| Number of Awakenings | 21 ± 9 | 17 ± 13 | 332.0, 0.151 |
| Wake After Sleep Onset (min) | 57.71 ± 45.11 | 39.64 ± 37.23 | 326.0, 0.265 |
| Stage N1 (%) | 4.63 ± 2.89 | 4.39 ± 4.52 | 320.0, 0.426 |
| Stage N2 (%) | 60.58 ± 9.00 | 61.11 ± 7.03 | 298.0, 0.750 |
| Stage N3 (%) | 14.00 ± 6.03 | 14.33 ± 5.64 | 301.0, 0.874 |
| Stage REM (%) | 20.79 ± 5.34 | 20.17 ± 3.89 | 312.0, 0.710 |

### TES^15kHz^-TI^1Hz^ preserves sleep architecture

Participants did not report feeling the stimulation during the night. In addition, we compared sleep architecture metrics across baseline, first, and last intervention nights for those TES^15kHz^-TI^1Hz^ participants with usable data on all nights (**Table 3**, TES^15kHz^-TI^1Hz^ N=16). There was a weak main effect of night on Total Sleep Time (X^2^=6.12, p=0.047) when assessed with a non-parametric, omnibus Friedman test; however, individual pairwise comparisons were not statistically significant with correction for multiple comparisons using the Benjamini-Yekutieli procedure (Baseline v. First: W=35.5, p=0.093; First v. Last: W=82.5, p=0.453; Baseline v. Last: W=36.0, p=0.098, Wilcoxon signed rank test). There were no differences in any other sleep characteristics when assessed with Friedmans tests or individual pairwise comparisons with correction for multiple comparisons. In TES^15kHz^ participants (**Supplementary Table 1**, TES^15kHz^ N=5), the Friedman test showed an overall effect of night on Sleep Onset Latency (X^2^=8.40, p=0.015); however, individual pairwise comparisons were not statistically significant with correction for multiple comparisons using the Benjamini-Yekutieli procedure (Baseline v. First: W=13.0, p=0.188; First v. Last: W=15.0, p=0.063; Baseline v. Last: W=15.0, p=0.063, Wilcoxon signed rank test). There were no differences in any other sleep characteristics. SWA (power spectral density, PSD in 0.5–4 Hz range) was quantified throughout each entire night then averaged across channels for each participant. There was no main effect of night (baseline, first, or last) on whole-night SWA in either the TES^15kHz^-TI^1Hz^ or TES^15kHz^ group (TES^15kHz^-TI^1Hz^: N=16, df=2, F=1.14, p=0.334; TES^15kHz^: N=5, df=2, F=3.65, p=0.075, one-way repeated measures ANOVAs). Similar results were found when SWA was measured during the first four hours after sleep onset; this early SWA did not differ between baseline, first, or last nights in either group (TES^15kHz^-TI^1Hz^: N=16, df=2, F=0.78, p=0.467; TES^15kHz^: N=5, df=2, F=3.07, p=0.103, one-way repeated measures ANOVAs). Additionally, the number of stimulation protocols completed during NREM sleep did not differ between first and last nights for either group (TES^15kHz^-TI^1Hz^: N=16, First=10±2, Last=9±1, W=29.0, p=0.910; TES^15kHz^: N=5, First=9±1, Last=8±2, W=13.0, p=0.250, Wilcoxon signed rank test).

**Table 3.** Sleep characteristics across nights in TES^15kHz^-TI^1Hz^. Values are Mean ± standard deviation.

|  | Baseline Night | First Night | Last Night | Test Statistic (X<sup>2</sup>), p-value |
| --- | --- | --- | --- | --- |
| Total Sleep-Wake Period (hr) | 8.06 ± 0.99 | 8.25 ± 0.85 | 8.12 ± 0.78 | 0.88, 0.646 |
| Total Sleep Time (hr) | 6.70 ± 0.83 | 7.04 ± 0.69 | 6.92 ± 0.79 | 6.12, 0.047<sup>†</sup> |
| Sleep Onset Latency (min) | 22.97 ± 11.89 | 23.44 ± 13.38 | 21.50 ± 15.39 | 2.51, 0.285 |
| Sleep Efficiency (%) | 83.62 ± 8.89 | 85.65 ± 6.65 | 85.33 ± 7.27 | 1.12, 0.570 |
| Number of Awakenings | 21 ± 7 | 21 ± 8 | 20 ± 8 | 1.76, 0.414 |
| Wake After Sleep Onset (min) | 58.19 ± 47.17 | 48.56 ± 36.60 | 50.13 ± 36.78 | 2.13, 0.345 |
| Stage N1 (%) | 4.07 ± 1.85 | 3.96 ± 2.94 | 4.07 ± 3.71 | 0.38, 0.829 |
| Stage N2 (%) | 61.49 ± 9.83 | 61.88 ± 5.76 | 60.03 ± 5.65 | 0.38, 0.829 |
| Stage N3 (%) | 13.75 ± 6.40 | 13.50 ± 6.35 | 12.62 ± 6.31 | 0.88, 0.646 |
| Stage REM (%) | 20.70 ± 5.73 | 20.66 ± 2.91 | 23.28 ± 3.79 | 1.62, 0.444 |

### SWA increases during and after TES^15kHz^-TI^1Hz^

We measured SWA in the immediate three-minutes before (PRE), during (STIM), and after (POST) TES^15kHz^-TI^1Hz^ stimulation and calculated the differences between time periods (**Figure 2A**). SWA was normalized (%SWA) to account for SWA differences between participants and night-to-night variability within participants (see Methods, hd-EEG Spectral Power Analyses). Since SWA changes were consistent on the first and last intervention nights (**Supplementary Figure 1**), we averaged Δ%SWA across the two nights for each participant. We also included participants (N=5) with only one intervention night of usable data to increase statistical power.

**Figure 2.**
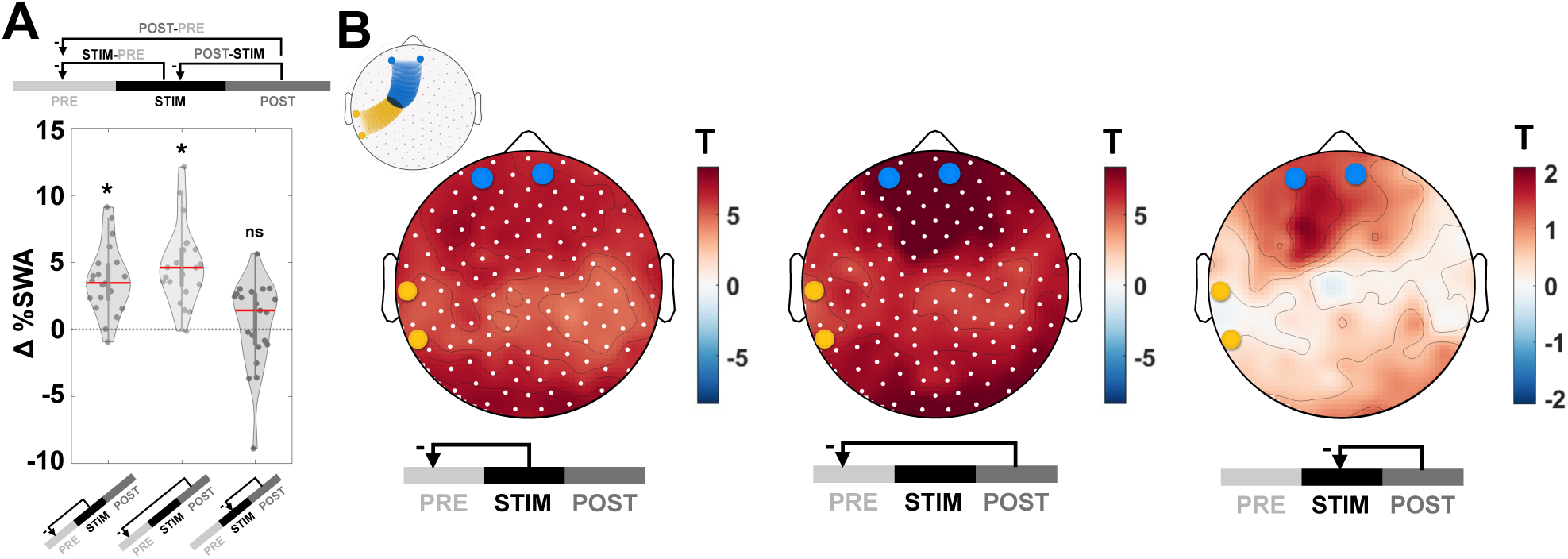
TES^15kHz^-TI^1Hz^ effects on SWA. **A**. Across all channels, normalized SWA (%SWA) increased during and after TES^15kHz^-TI^1Hz^ compared to before. %SWA did not change after stimulation compared to during stimulation. Two-tailed paired samples t-tests used to evaluate. Median shown as solid red line. **B**. Increases in %SWA during and after stimulation were global. No channels had statistically significant changes in %SWA after stimulation compared to during stimulation. Topography of T-statistic shown; channels with statistically significant changes are indicated by white dots. Approximate locations of stimulating electrodes are shown in blue (E_1_) and yellow (E_2_). See Supplementary Table 3 for details on figure statistics.

Overall, we found a significant difference in the average SWA across PRE, STIM, and POST periods (N=21, df=2, F=30.97, p=7.48×10^-9^, one-way repeated measures ANOVA). We found a global (across all channels) increase in SWA during the stimulation as compared to before stimulation (**Figure 2A**, STIM-PRE, N=21, t=6.64, p=1.81×10^-6^, d=1.45; two-tailed paired samples t-test). A global increase in SWA was also present after stimulation compared to before stimulation (**Figure 2A**, POST-PRE, N=21, t=7.47, p=3.34×10^-7^, d=1.63; two-tailed paired samples t-test). There were no significant differences between the periods after (POST) stimulation and those during (STIM) stimulation in the average across all channels (**Figure 2A**, POST-STIM, N=21, t=0.63, p=0.536, d=0.14, two-tailed paired samples t-test). To identify statistically significant topographic changes at the level of individual channels, we implemented a nonparametric cluster-based statistical approach using a suprathreshold cluster analysis to control for multiple comparisons. The results of this topographical analysis confirmed the results at the global level, i.e. SWA increased across all channels during stimulation and the increase persisted to a similar degree after, with no significant differences between the STIM and POST periods, despite a higher T-statistic in frontal regions in the POST period (**Figure 2B**, STIM-PRE: 1 positive cluster, 185 significant electrodes, p̄=1.93×10^-5^; POST-PRE: 1 positive cluster, 185 significant electrodes, p̄=6.87×10^-6^; POST-STIM: 0 clusters, 0 significant electrodes, p̄=0.537).

### TES^15kHz^-TI^1Hz^ effects on SWA differ from those of TES^15kHz^

We then performed a preliminary comparison of the effects on SWA of TES with amplitude modulation (TES^15kHz^-TI^1Hz^, 1 Hz difference) and without (TES^15kHz^, 0 Hz difference) (**Figure 3A**). To do so, we directly compared normalized SWA (%SWA) in the STIM period between TES^15kHz^-TI^1Hz^ and TES^15kHz^. First, we used nonparametric cluster-based statistics to identify channels with significantly different SWA during stimulation between TES^15kHz^-TI^1Hz^ and TES^15kHz^ (1 positive cluster, 107 significant electrodes, p̄=0.028), and then we calculated the average SWA across these channels. We found that, during STIM, SWA in TES^15kHz^-TI^1Hz^ was greater than SWA in TES^15kHz^ (**Figure 3B**, TES^15kHz^-TI^1Hz^ N=21, TES^15kHz^ N=7, t=2.48, p=0.020, d=1.08, We also tested whether the effects on SWA changed throughout the STIM period and, if so, whether these changes occurred both with TES^15kHz^-TI^1Hz^ and TES^15kHz^. For this analysis, we compared the last 45 seconds (LATE) of the STIM period with the first 45 seconds (EARLY), including the 15-second ramping interval (**Figure 3C**). In the TES^15kHz^-TI^1Hz^ group, SWA was higher in LATE compared to EARLY (**Figure 3D**, N=21, t=3.25, p=4.02×10^-3^, d=0.71; two-tailed paired samples t-test). Increases in SWA in LATE were global and strongest in the frontal region (**Figure 3E**, 1 positive cluster, 165 significant electrodes, p̄=8.83×10^-3^). By contrast, in TES^15kHz^, SWA did not change between LATE and EARLY (**Figure 3D**, N=7, t=1.71, p=0.138, d=0.65; two-tailed paired samples t-test, **3E,** 0 clusters, 0 significant electrodes, p̄=0.200).

**Figure 3.**
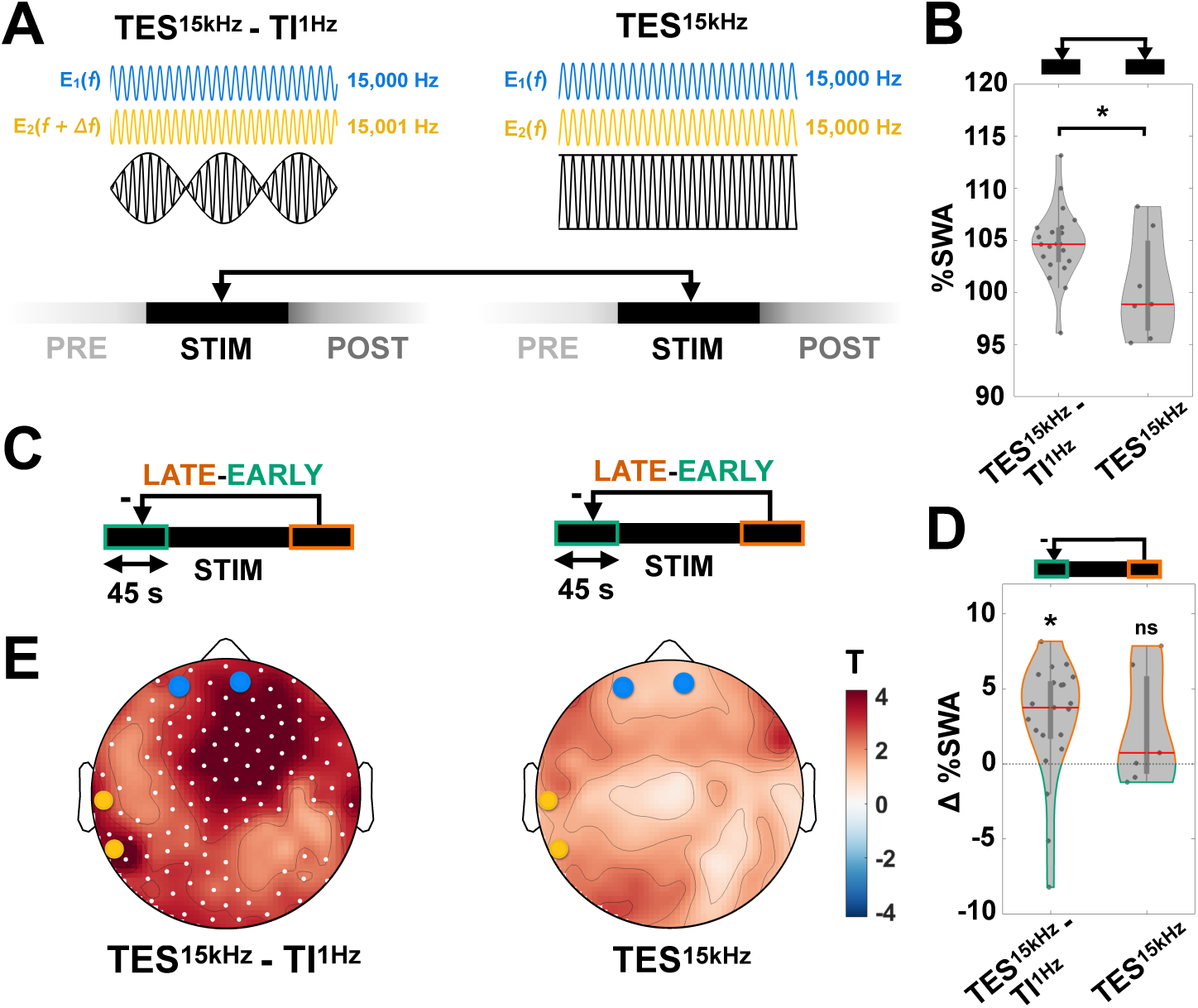
TES^15kHz^-TI^1Hz^ versus TES^15kHz^ effects. **A**. In TES^15kHz^-TI^1Hz^, one pair of electrodes delivers high frequency alternating current with an offset (15,001 Hz) to create temporal interference (amplitude modulation) at 1 Hz. In TES^15kHz^, both pairs of electrodes deliver the same high frequency alternating current, with no offset, i.e. there is no amplitude modulation (difference is 0 Hz). Normalized SWA (%SWA) during the stimulation period (STIM) was directly compared between TES^15kHz^-TI^1Hz^ and TES^15kHz^. **B**. Average %SWA across channels with statistically significant differences between TES^15kHz^-TI^1Hz^ and TES^15kHz^ (1 positive cluster, 107 significant electrodes, p!=0.028) are shown. %SWA was higher during TES^15kHz^-TI^1Hz^ as compared to TES^15kHz^ (two-tailed independent samples t-test). Median shown as solid red line. **C**. Within the three-minute period during stimulation, early and late forty-five second intervals were considered; the difference in %SWA between the late and early stimulation intervals was compared for both TES^15kHz^-TI^1Hz^ and TES^15kHz^. **D**. Averaged across all channels, %SWA was greater in the late interval compared to the early interval in TES^15kHz^-TI^1Hz^ but not in TES^15kHz^. Two-tailed paired samples t-tests were used. **E**. (Left) In TES^15kHz^-TI^1Hz^, increases in %SWA later in stimulation were global. (Right) In TES^15kHz^, no channels had statistically significant changes in %SWA between late and early intervals. Topography of T-statistic shown; channels with statistically significant changes are indicated by white dots. Approximate locations of stimulating electrodes are shown in blue (E_1_) and yellow (E_2_). See Supplementary Table 3 for details on figure statistics.

### TES^15kHz^-TI^1Hz^ effects on other frequency bands

Next, we tested whether TES^15kHz^-TI^1Hz^ had additional effects on the EEG power spectrum. PSD values were averaged for each PRE, STIM, and POST period, then across electrodes, then across first and last intervention night, to yield one spectrum per participant (N=21). Spectra were then averaged across participants (**Figure 4**). For each frequency bin (N=240, 0.167 Hz steps), the STIM-PRE difference, as well as the POST-PRE difference, were calculated. For statistical comparisons, for each canonical band (e.g., theta, alpha), PSD values were averaged across the frequencies in that band for each participant, then STIM-PRE and POST-PRE comparisons were performed using paired samples t-tests with the Benjamini-Yekutieli procedure to correct for multiple comparisons (**Supplementary Table 2**). For selected significant topographical analyses, as was done with SWA, band power was normalized (% band power) by the average band power during NREM throughout the entire night to give a relative change from the mean overnight band power. Then, the difference between time periods (STIM-PRE, POST-PRE) was evaluated with nonparametric cluster-based statistics.

**Figure 4.**
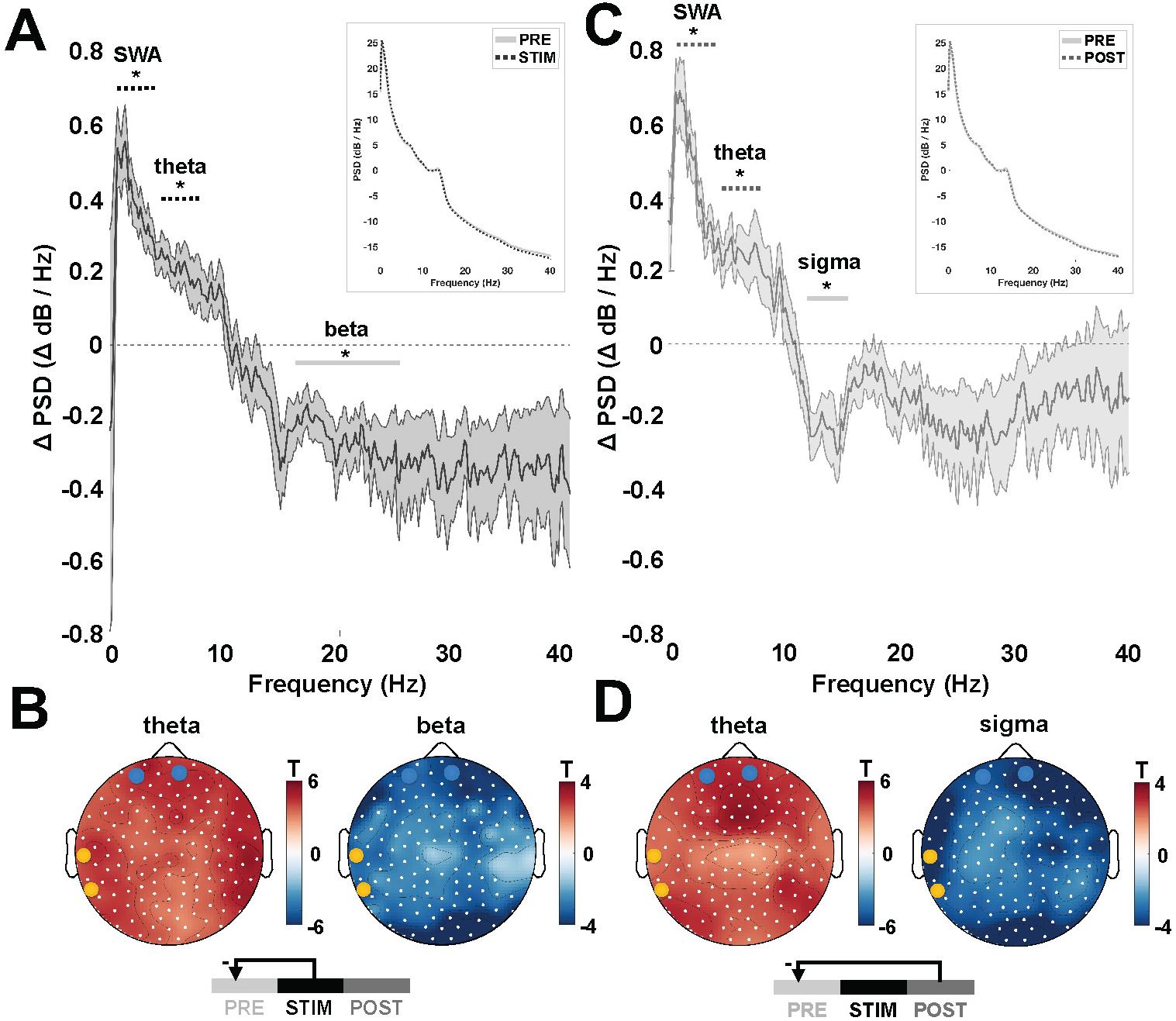
TES^15kHz^-TI^1Hz^ effects on other frequency bands. **A**. Differences in power spectral density (PSD) during (STIM) compared to before (PRE) stimulation for individual frequencies (N=240, 0.167 Hz steps). PSD was averaged across channels then across participants (dark line) with the standard error of the mean across participants (light lines) shown. In addition to SWA (Figure 2), increased theta and decreased beta power were found during stimulation. Two-tailed paired samples t-tests were used with correction for multiple comparisons across bands using the Benjamini-Yekutieli procedure. (Top Right, Inset) PSD for individual frequencies before (PRE) and during (STIM) stimulation. **B**. Changes in normalized theta and normalized beta power were global. Topography of T-statistic shown; channels with statistically significant changes are indicated by white dots. Approximate locations of stimulating electrodes are shown in blue (E_1_) and yellow (E_2_). **C**. Differences in PSD after (POST) compared to before (PRE) stimulation. In addition to SWA (Figure 2), increased theta and decreased sigma power were found after stimulation. (Top Right, Inset) PSD for individual frequencies before (PRE) and after (POST) stimulation. **D**. Changes in normalized theta and normalized sigma power were global. See Supplementary Table 3 for details on figure statistics.

We found that, as compared to before, during stimulation, TES^15kHz^-TI^1Hz^ increased power in the theta (4–8 Hz) range and decreased power in the beta (16–25 Hz) range (**Figure 4A**, STIM-PRE, N=21, theta: t=4.29, p=3.58×10^-4^, d=0.94; beta: t= -3.29, p=3.68×10^-3^, d= -0.72; two-tailed paired samples t-tests). Both changes were global (**Figure 4B**, theta: 1 positive cluster, 185 significant electrodes, p̄=9.07×10^-4^; beta: 1 negative cluster, 173 significant electrodes, p̄=7.59×10^-3^). As compared to before stimulation, after TES^15kHz^-TI^1Hz^, we found an increase in power in the theta range and a decrease in the sigma (12–16 Hz) range (**Figure 4C**, POST-PRE, N=21, theta: t=3.90, p=8.92×10^-4^, d=0.85; sigma: t= -3.95, p=7.95×10^-4^, d= -0.86; two-tailed paired samples t-tests). All these changes were also global (**Figure 4D**, theta: 1 positive cluster, 185 significant electrodes, p̄=1.46×10^-3^; sigma: 1 negative cluster, 185 significant electrodes, p̄=4.47×10^-3^).

When the analysis was repeated for TES^15kHz^, there were no differences in any frequency band for either the STIM-PRE or POST-PRE comparisons (N=7, STIM-PRE: SWA: t=1.34, p=0.229; theta: t=0.98, p=0.366; alpha: t= -0.75, p=0.482; low sigma: t= -0.98, p=0.364; high sigma: t= -1.42, p=0.205; beta: t= -0.46, p=0.660; gamma: t= -0.97, p=0.367; POST-PRE: SWA: t=1.23, p=0.264; theta: t=1.98, p=0.095; alpha: t= -0.74, p=0.487; low sigma: t= -0.93, p=0.387; high sigma: t= -2.67, p=0.037 (adjusted p=1.676); beta: t= -0.79, p=0.459; gamma: t= -1.55, p=0.172).

### Changes in SWA during TES^15kHz^-TI^1Hz^ are associated with improved sleep quality ratings following the TES^15kHz^-TI^1Hz^ intervention

Finally, we examined potential relationships between stimulation-related changes in normalized SWA (%SWA) and participants’ perceptions of how they slept. For each participant in the TES^15kHz^-TI^1Hz^ group, we compared the difference in SWA during (STIM-PRE) and after (POST-PRE) TES^15kHz^-TI^1Hz^ with subjective sleep quality ratings from the Restorative Sleep Questionnaire (REST-Q) ^29,30^, assessed the morning after each intervention night. In line with Robbins and colleagues ^29^, an overall REST-Q score was calculated for each participant and each intervention night. When evaluating first and last night separately, using Spearman’s correlations (r_s_), we found no correlations between sleep quality ratings and SWA during or after TES^15kHz^-TI^1Hz^ on either night (STIM-PRE, N=16; First, r_s_=0.038, p=0.888; Last, r_s_=0.159, p=0.555; POST-PRE, N=16; First, r_s_=0.176, p=0.515; Last, r_s_= -0.218, p=0.416). We then considered how SWA and sleep quality ratings might change over the course of the intervention, from first to last night, in the context of inter-individual variability. We found that a greater increase in SWA during TES^15kHz^-TI^1Hz^ (STIM-PRE) on the last night compared to the first night was associated with higher overall REST-Q scores on the last night compared to the first night (**Figure 5A**, Left, N=16, r_s_=0.563, p=0.023). This association was strongest for channels over frontal regions (**Figure 5A**, Right, 2 positive clusters, 73 significant electrodes, p̄=0.016). To examine the specificity of the association found for SWA, we also tested the relationship between the change in normalized sigma power and the change in overall REST-Q scores from first to last intervention night, and found no significant association for either low or high sigma power (**Figure 5B**, Low Sigma: 9–12 Hz, N=16, r_s_=0.152, p=0.573; Right: 0 clusters, 0 significant electrodes, p̄=0.549; **Figure 5C**, High Sigma: 12–16 Hz, N=16, r_s_= -0.229, p=0.393; Right: 0 clusters, 0 significant electrodes, p̄=0.597).

**Figure 5.**
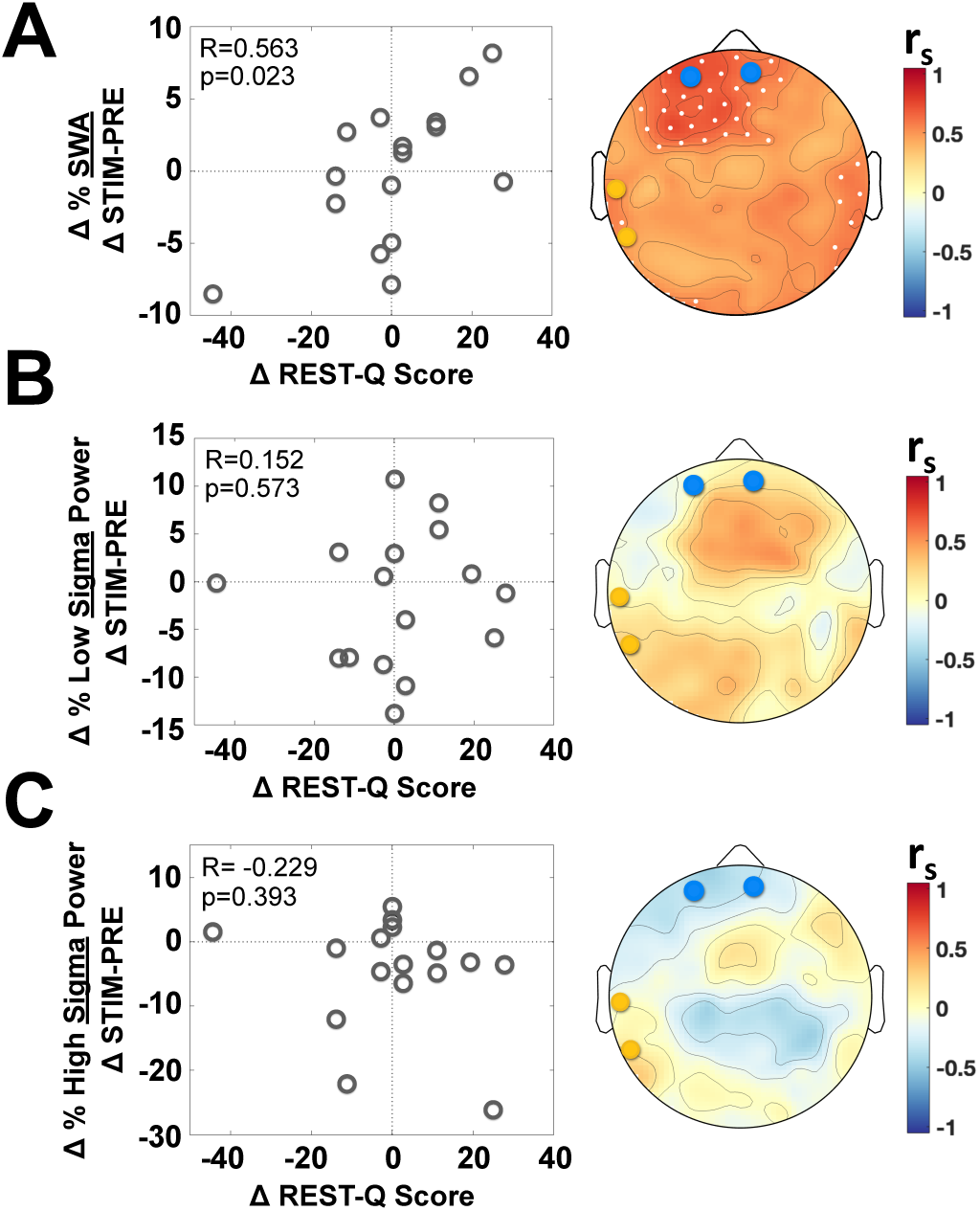
Relationship between changes in SWA during TES^15kHz^-TI^1Hz^ and changes in sleep quality following TES^15kHz^-TI^1Hz^ intervention. Greater increases in normalized SWA (%SWA), averaged across all channels, were related to improved sleep quality in TES^15kHz^-TI^1Hz^. **A**. (Left) Difference in %SWA during stimulation (STIM-PRE, y-axis) was positively correlated with the difference in overall REST-Q scores on the last intervention night as compared to the first intervention night (x-axis). (Right) Strongest correlations were present in frontal areas. Topography of Spearman’s correlation coefficients are shown; channels with statistically significant changes are indicated by white dots. Approximate locations of stimulating electrodes are shown in blue (E_1_) and yellow (E_2_). **B**. (Left) Difference in normalized 9–12 Hz (low sigma) power during stimulation (STIM-PRE, y-axis) was not correlated with the difference in overall REST-Q scores on the last intervention night as compared to the first intervention night (x-axis). (Right) No channels had statistically significant correlations. **C**. (Left) Difference in normalized 12–16 Hz (high sigma) power during stimulation (STIM-PRE, y-axis) was not correlated with the difference in overall REST-Q scores on the last intervention night as compared to the first intervention night (x-axis). (Right) No channels had statistically significant correlations. See Supplementary Table 3 for details on figure statistics.

## Discussion

We performed simultaneous hd-EEG recording and Transcranial Electrical Stimulation with Temporal Interference (TES-TI, 15 kHz, 1 Hz difference) during overnight sleep in a laboratory setting in healthy humans with the goal of enhancing SWA during NREM sleep. The results show that TES^15kHz^-TI^1Hz^ targeting left ventromedial prefrontal cortical regions increased SWA during the stimulation period, and the effect outlasted the stimulation. TES^15kHz^-TI^1Hz^ during the stimulation period also increased SWA compared to TES^15kHz^ without TI (15 kHz, 0 Hz difference), indicating that the effect is specifically attributable to TI. Moreover, increases in SWA were greater in the late part of the stimulation interval with TES^15kHz^-TI^1Hz^ stimulation but not with TES^15kHz^. Effects on spectral power in other frequency bands were also found, pointing to an overall increase in low frequencies (SWA and theta range) and a decrease in higher frequencies (sigma and beta range) during and/or after stimulation. Lastly, participants who showed the largest incremental effects on SWA between the first and the last intervention night of the 4-week protocol also showed the largest improvement in restorative sleep ratings. Together, these findings illustrate the ability of TES^15kHz^-TI^1Hz^ to enhance SWA and, more generally, the potential for TES-TI as an intervention to improve sleep.

TES-TI offers several potential advantages compared to other techniques for neuromodulation during sleep. While acoustic stimulation increases SWA ^31,32^, the closed-loop titration of the intensity of stimuli to effectively trigger slow waves without awakening subjects can be problematic. This is likely because the triggering of slow waves is mediated by the non-lemniscal auditory pathway, which is associated with the ascending reticular activating system (ARAS) in the brainstem ^10^. By directly modulating slow wave generation within their primary cortical origination sites, TES-TI circumvents unintended ARAS activation. Indeed, when comparing baseline, first and last intervention nights, we found that TES^15kHz^-TI^1Hz^ did not affect the general sleep architecture and, more specifically, did not increase the number of awakenings and the time spent awake after sleep onset (Table 3). Notably, the percent of time spent in slow wave sleep (Stage N3) and SWA (both throughout the entire night and in the first four hours of sleep) did not change across baseline, first, and last intervention nights for TES-TI participants. This is in line with several studies using an ON/OFF block design to enhance SWA with other methods^14^. It is possible that more numerous stimulation protocols and/or continuous stimulation might expand the increases in SWA observed in the immediate 3-minutes during and after TES-TI to a larger timescale, over several hours. Also consistent with the preservation of sleep architecture was that the power spectra before, during, and after stimulation all maintained a profile consistent with NREM sleep, including a large, broad peak in the low frequencies corresponding to SWA and a smaller peak in the higher frequencies corresponding to sigma power.

Conventional TES, which typically employs frequencies below 1 kHz, has also been used to increase sleep SWA ^12,33,34^. However, conventional TES is not well suited for reaching deep brain regions and to do so focally, it produces scalp discomfort and potential awakenings at relatively low stimulation intensities (e.g., 1.5 mA). Also, electrical artifacts prevent the analysis of concurrent EEG recordings ^14^. By contrast, TES^15kHz^-TI^1Hz^ as performed in this study, can reach deep brain regions with relatively high focality and field intensity, as indicated by electric field modeling for individual subjects. Even at the relatively high intensities employed in this study (5 mA), TES^15kHz^-TI^1Hz^ did not produce any scalp sensation and did not awaken subjects. In addition, given the high carrier frequency (15 kHz, much higher than the EEG signal) and the use of dedicated hardware filters, it was possible to conduct simultaneous TES^15kHz^-TI^1Hz^ and hd-EEG recordings avoiding stimulation artifacts (there were no narrow peaks at 1Hz or its harmonics superimposed on the smooth power spectra typical of NREM sleep).

TES^15kHz^-TI^1Hz^ was intentionally targeted to a deep brain region—left ventromedial prefrontal cortical regions that we had previously identified as a “hot spot” for the origination of slow waves through source modeling of all-night hd-EEG recordings ^27^. Because slow waves travel across the cortical surface ^35^ we nevertheless suspected that the effects of targeted neuromodulation might extend to broader cortical areas. In fact, the increases in SWA during and after TES^15kHz^-TI^1Hz^ were global, extending across all channels.

An interesting aspect of the present results is that the increase in SWA produced by TES^15kHz^-TI^1Hz^ was incremental—increasing from the beginning to the end of the 3-minute stimulation period. It also outlasted the stimulation, extending to the subsequent 3 minutes. Because of the experimental design, we could not properly assess how long the enhancement may have lasted. Nevertheless, these findings suggest that TES^15kHz^-TI^1Hz^ at 1 Hz may not simply boost, synchronize, or entrain neuronal slow oscillations, but may produce plastic changes that underlie a cumulative effect. These findings clearly warrant further investigation of stimulation protocols that may maximize the effectiveness of SWA enhancement by lengthening stimulation periods, increasing the number of stimulation periods, and/or repeating them over subsequent nights.

An important part of this study was the direct comparison of TES^15kHz^-TI^1Hz^ with TES^15kHz^. A separate group of participants (N=7) received high frequency stimulation (15 kHz) with the same intensity and duration as the TES^15kHz^-TI^1Hz^ group, but with 0 Hz frequency difference between the two stimulation channels (TES^15kHz^). High frequency TES is often assumed to produce minimal effects on the brain because of dampening of the field intensity by the skull, resulting in subthreshold intensities, but this assumption has not yet been tested fully ^22,36^. The present results showed that TES^15kHz^-TI^1Hz^, compared to TES^15kHz^, enhanced SWA during the stimulation period. Moreover, SWA was higher in the last 45 seconds of stimulation compared to the first 45 seconds during TES^15kHz^-TI^1Hz^ and no different during TES^15kHz^. Although there were fewer TES^15kHz^ participants, these initial findings support the notion that the focal amplitude modulation at 1 Hz obtained through TES^15kHz^-TI^1Hz^ has specific effects on brain activity that are not observed by mere high frequency TES^15kHz^, consistent with data from intracranial recordings ^22^.

Besides enhancing sleep SWA, the TES^15kHz^-TI^1Hz^ intervention employed here produced changes in the sleep power spectrum that were consistent with an overall deepening of sleep. In addition to increased power in the SWA (delta) band, we observed a global increase in theta power during and after TES^15kHz^-TI^1Hz^ stimulation. Sleep onset is typically accompanied by an increase in both delta and theta power ^37,38^ and recovery sleep is characterized by increases in low frequency power that extend into the theta band ^39^. Similarly, the global decrease in beta power concomitant with increased SWA during TES^15kHz^-TI^1Hz^ was consistent with the inverse relationship between beta and delta power and the higher delta:beta ratio associated with reduced arousal levels ^38,40^. The decrease in power in the sigma range (12–16 Hz) after TES^15kHz^-TI^1Hz^ (and a trend during TES^15kHz^-TI^1Hz^) may also reflect a deepening of sleep. In general, a reduction in spindles is seen with the transition to slow wave sleep (Stage N3), in recovery sleep after total sleep deprivation ^39^ and, in particular, fast spindle power has dynamics that run in the opposite direction of those of SWA ^41,42^.

A promising indication of this study was the improvement in the restorative quality of sleep when comparing the first and last intervention nights of TES^15kHz^-TI^1Hz^, as reported by subjects through REST-Q scores. Specifically, participants with greater increases in SWA during TES^15kHz^-TI^1Hz^ from the first to the last night also reported better overall restorative sleep ratings. This relationship was specific to sleep SWA, as it did not extend to the sigma range. The ability of TES^15kHz^-TI^1Hz^ to produce effects on neuronal activity during sleep that outlast the stimulation and may be associated with plastic changes, as identified here, will be further investigated in the STRENGTHEN study by assessing long-lasting modifications in brain anatomical and functional connectivity, as well as in behavioral indicators of cognitive flexibility and emotional regulation ^43,44^.

This first study of TES-TI during sleep had several limitations. We targeted left ventromedial prefrontal cortex using a standardized montage for TES^15kHz^-TI^1Hz^. Future work may determine whether more precise, individualized TES-TI targeting may offer superior efficacy ^45^. Our initial analyses focused on the effects of TES^15kHz^-TI^1Hz^ during NREM sleep on first and last intervention nights. Future work within the STRENGHTEN framework will evaluate additional intervention nights as well as the effects of TES^15kHz^-TI^6Hz^ during REM sleep. Further enrollment will increase the number of subjects receiving TES^15kHz^ and possibly add a within-participant component. On the other hand, few clinical trials of TES-TI have provided a direct comparison with TES ^22^, as was done here.

SWA varies with age, and the age range in this study is relatively wide (19–49 years). However, we assessed the effect of TES^15kHz^-TI^1Hz^ with an intra-subject design, by comparing adjacent 3-minute periods and normalizing power to the entire night ^38,46-48^. While fluctuations of SWA during NREM episodes can present a challenge, the use of multiple stimulation blocks (repeated ten times per night) primarily throughout the first four hours of NREM sleep was intended to avoid spurious effects associated with infra-slow oscillations or the natural deepening of sleep. Future studies will aim at identifying the optimal parameters for TES-TI stimulation, including duration and timing, informed by increasing knowledge about its mechanism of action ^18,25,49^. Moreover, the ability of TES-TI to modulate other features of sleep, from sleep spindles to sawtooth waves to sharp-wave ripples, are also ready for exploration, with potential applications in health and disease.

## Methods

### Study Design

Data used in this study were acquired as part of the STRENGTHEN clinical trial (NCT06267521), a single-blind, longitudinal study aimed at enhancing cognitive flexibility and emotion regulation. The study design includes Transcranial Electrical Stimulation with Temporal Interference (TES-TI) delivered during sleep in the lab as well as brief, daily meditation training. Healthy participants (ages 18–50) were assigned to 1 of 4 groups: (Group 1) TES^15kHz^ (0 Hz difference frequency) 2 night per week and meditation; (Group 2) TES^15kHz^-TI^1Hz^ (1 Hz difference frequency) 2 nights per week and meditation; (Group 3) TES^15kHz^-TI^1Hz^ 1 night per week and meditation; (Group 4) TES^15kHz^-TI^1Hz^ 2 nights per week and sham meditation. For all participants, interventions during sleep in the lab occurred over four consecutive weeks. Participants were assigned to their treatment group following baseline assessment, with efforts made to ensure group equivalence on key demographic variables, particularly age and sex. This matching approach mitigated potential confounds related to sex and aged-based differences in sleep. Participants were blind to experimental group, i.e. they did not know whether they were receiving TES^15kHz^ (Group 1) or TES^15kHz^-TI^1Hz^ (Groups 2,3,4). Participants were reimbursed for their study participation. All aspects of the study were approved by University of Wisconsin-Madison Institutional Review Board.

The current study reflects a mechanistic sub-study within the broader STRENGTHEN trial; it focuses on the effects of TES^15kHz^-TI^1Hz^ during NREM sleep using data collected from the first five cohorts (out of nine) of the STRENGTHEN trial. The analyses presented here were not pre-specified in the formal clinical trial registration; however, they were part of the planned exploratory aims within the Institutional Review Board-approved protocol and were designed prior to data collection from the analyzed cohorts. Primary and secondary endpoints of the trial related to cognitive flexibility and emotion regulation will be the focus of later work. The analyses performed here were initiated after interim inspection of data from the first two cohorts showed large effect sizes. Specifically, analyses, as in Figure 2, performed after the first two cohorts (N=10 TES^15kHz^-TI^1Hz^) indicated effect sizes ranging 0.947–1.146 (Cohen’s d) suggesting 5–9 subjects would be sufficient to obtain statistical power of 0.8 (calculated using MATLAB *sampsizepwr* for a one-tailed paired samples t-test). To maximize statistical power, the current study used data from the first five cohorts to examine the effects of TES^15kHz^-TI^1Hz^ on NREM sleep (N=21, 1 Hz, targeting left ventromedial prefrontal cortex during NREM sleep episodes in the first half of the night) by comparing the baseline night with the first and last intervention nights regardless of the group. A preliminary comparison between TES^15kHz^-TI^1Hz^ and TES^15kHz^ (i.e., directly assessing the potential effects of amplitude modulation) was also performed utilizing Group 1 participants from the first five cohorts (N=7). Another goal of the STRENGTHEN clinical trial, to be reported in a subsequent publication, was the assessment of TES^15kHz^-TI^6Hz^ (6 Hz, targeting retrosplenial cortex) during REM sleep episodes later in the night.

### Participants

In total, twenty-eight participants (30.5±9.9 years, 61.0% Female) from the first five cohorts of the STRENGTHEN study were included in the current study. All participants were medically healthy and free of sleep disorders. Participants had a baseline magnetic resonance imaging (MRI) scan and sleep night in the lab. The baseline sleep night did not include overnight stimulation. Participants were excluded if they had any neuroradiologist-identified structural abnormalities of the brain. Participant recruitment began with an online survey. Potential participants were then further screened by phone call to see if they met eligibility criteria. They then underwent additional screening and an informed consent discussion in person prior to enrollment in the study. All participants gave written informed consent in accordance with the University of Wisconsin-Madison Institutional Review Board.

### Datasets Included

Baseline, first, and last intervention nights were utilized here; a future publication will report on all the intervention nights. In total, twenty-two first nights and twenty-seven last nights were included in subsequent analyses that evaluated effects during stimulation. Three first nights were excluded due to technical issues and two first nights were excluded due to poor sleep that precluded experiment completion (e.g., fewer than five out of ten intended stimulation protocols were completed). For two additional nights (one first and one last), hd-EEG recordings during the stimulation were excluded due to technical issues. However, these two nights were included in subsequent analyses that evaluated effects before or after stimulation. Analyses comparing baseline, first, and/or last intervention nights were restricted to participants with usable data across all nights.

### Procedure

Participants spent overnights in the sleep lab at the Wisconsin Sleep Center, where they were outfitted with a 256-channel hd-EEG net for recording brain activity and additional channels for recording polysomnography (PSG) data (electrooculography adjacent to lateral canthi, submental electromyography, and electrocardiography). Channels for performing TES-TI were embedded in the hd-EEG net, placed in lieu of recording channels. While the participant was awake, short stimulation protocols (15-seconds with 15-second ramping up/down periods) were performed with increasing current amplitudes (up to 5 mA peak to peak) to determine if a participant began to feel sensation on their scalp. The current amplitude was then lowered systematically and checked again such that the final current value of 5 mA chosen did not elicit any sensation. Then, the participant went to sleep, and a sleep technician performed stimulation protocols based on sleep features visible on the hd-EEG and PSG recordings. Specifically, when NREM sleep was identified and stable (≥3 minutes), stimulation was delivered in 3-minute protocols with 15-second ramping up/down periods. Each protocol was separated by at least 6 minutes and repeated approximately ten times, in most cases confined to NREM sleep episodes during the first four hours after sleep onset. Participants were monitored throughout the night from a control room and a sleep technician was available for communication at any time.

### Sleep Quality Surveys

After each overnight in the sleep lab, participants completed the 9-question Restorative Sleep Questionnaire (REST-Q) survey ^29,30^. They rated to what extent they felt tired, sleepy, in a good mood, rested, refreshed, ready to start the day, energetic, mentally alert, and grouchy on scales of 1 to 5 (1=Not at all, 2=A little bit, 3=Some, 4=Very much, 5=Completely). An overall REST-Q score was calculated in line with ^29^. Overall scores from after the first and last intervention nights were included in the analyses. Potential associations between scores and changes in SWA on either the first or last intervention night were evaluated using Spearman’s correlations. Then, associations between the change in scores and the change in band power from first to last intervention night were also evaluated using Spearman’s correlations. Specifically, the difference between overall REST-Q scores on the last intervention night compared to the first intervention night was computed. The difference between SWA (0.5–4 Hz) and sigma (low 9–12 Hz, high 12–16 Hz) band power on the last intervention night compared to the first was computed. Potential relationships between these differences were then assessed with Spearman’s correlations.

### TES-TI Targeting

A standardized channel montage was used for all participants (channel 1: E20–E34, channel 2: E70–E95 of the hd-EEG net) targeting left ventromedial prefrontal cortical regions. Channel placement was first approximated using the Temporal Interference Planning Tool (TIP v1.0, IT’IS Foundation, Switzerland), in which an optimal channel configuration was chosen from the 10:10 system to target the left ventromedial prefrontal cortex with maximal intensity and minimal off-target stimulation ^50,51^. TIP simulations were performed using the comprehensive MIDA head model, which details the electrical properties for different tissues in a twenty-nine-year-old healthy female volunteer ^52^. For further confirmation, targeting was then verified using SimNIBS software (v4.1.0) that provided tools for electric field modeling with additional details including: an individual’s MRI, and channel material, geometry, and position in the 256 hd-EEG system ^53,54^. Modeling began with segmentation of an individual participant’s T1 and T2 scans to extract electrical properties of different tissues. Segmentation and meshing were performed using the *charm* function. The participant’s head model was then aligned to a template 256-channel hd-EEG net. Next, electric fields were simulated accounting for channel properties (see *TES-TI Stimulation*), a current value of 5 mA peak to peak, and location in the 256-channel hd-EEG net extrapolated from the 10:10 system locations used in TIP simulations. Simulation of the temporal interference field (V/m) was implemented using the function *get_maxTI* that calculates the maximal modulation amplitude of the difference frequency using the equation described in ^18^. The field values obtained in grey matter were then extracted and plotted over the individual’s T1 scan as well as over a standardized atlas (MNI152) for visualization using SimNIBS and AFNI (Analysis of Functional NeuroImages) software ^55,56^.

Overall, simulations performed in SimNIBS with individualized head models agreed with those performed in TIP using the standardized MIDA head model. Based on these simulations using individualized head models, across all TES^15kHz^-TI^1Hz^ participants (N=21), maximal intensity values in left ventromedial prefrontal cortex grey matter were 0.82±0.09 V/m, ranging 0.62–0.99 V/m and average intensity values were 0.46±0.05 V/m, ranging 0.36–0.52 V/m. Of note, the channel configuration used for two TES^15kHz^-TI^1Hz^ participants differed by one channel (E94 instead of E95); however, their data was included given that electric field simulations for both configurations were highly similar.

### TES-TI

Four silver-silver chloride stimulating channels (12.6 mm outer diameter, 8.0 mm inner diameter, 2.2 mm width) with 1.5-meter cable length were embedded into the hd-EEG net, attached to the participant’s scalp with conductive paste (Ten20), and then connected to the TI Brain Stimulator for Research (TIBS-R v3.0, TI Solutions AG, Switzerland). Electrode impedances were monitored throughout the experiment and served as an indicator for the need to adjust electrodes; substantial increases in impedance indicated poor positioning and a need for adjustment. Average impedances were 4590±110 Ohm for channel 1 and 5020±600 Ohm for channel 2. TES-TI protocols during NREM sleep had the following parameters: 15,000 Hz carrier frequency, 1 Hz difference frequency, sinusoidal waveform, 5 mA peak to peak, 3-minute duration with 15-second ramping up/down periods. Channel 1 used a 15,000 Hz frequency and channel 2 used a 15,001 Hz frequency. Actual, average current delivered was 5.21±0.21 mA for channel 1 and 5.06±0.19 mA for channel 2 as reported by the TIBS-R device. Programming of the TIBS-R device was performed using its Python software.

### hd-EEG Data Acquisition

The hd-EEG system from MagStim Electrical Geodesics, Inc. (EGI) consisted of a hd-EEG net, an amplifier, and a physiological input box. Each net had 256 silver-silver chloride electrodes and was appropriately sized to each individual participant. In cases where the stimulating electrodes were very close to the hd-EEG recording electrodes, e.g., small net sizes, surrounding recording electrodes were not gelled to avoid the possibility of bridging between stimulating and recording electrodes. Nets were then connected to EGI amplifiers that interface with EGI’s Net Station Acquisition software (v5.4.2) for real-time data visualization. The physiological input box was used to collect polysomnography data (electrooculography, electromyography, and electrocardiography). In addition, data from the TIBS-R trigger was input into the physiological input box. The optical trigger box output a 5 V square wave signal with the onset and offset of stimulation and, when synchronized to hd-EEG recordings, allowed for offline identification of stimulation intervals. All data were collected at a 500 Hz sampling rate.

Hardware filters specifically designed for the TIBS-R device allowed for simultaneous TES-TI and hd-EEG recording. A low-pass filter (7th order, 100 Hz cutoff) was placed in the path of the hd-EEG amplifier input such that signals from the hd-EEG net were filtered before they reached the amplifier. A high-pass filter (4th order, 250 Hz cutoff) was placed in the path of the TIBS-R output such that intermodulation products and low-frequency noise generated by the TIBS-R were filtered before they reached the stimulating electrodes. Both hardware filters were used to allow for simultaneous TES-TI stimulation and hd-EEG recording.

### MRI Data Acquisition

MRI data were collected using a 3 Tesla MAGNUS (Microstructure Anatomy Gradient for Neuroimaging with Ultrafast Scanning, GE Healthcare) head-only MRI scanner. Structural images were acquired using T1- and T2-weighted images, with 0.8 mm isotropic voxels, and the following parameters. T1-weighted: sequence = MP-RAGE (Magnetization-Prepared Rapid Gradient-Echo), repetition time (TR) = 2000 ms, echo time (TE) = 3 ms, inverse time (TI) = 1100 ms, flip angle = 8 degrees, field of view (FOV) = 256 x 256 mm^2^, matrix size = 320 x 320 pixels, resolution = 0.8 mm x 0.8 mm x 0.8 mm, number of slices = 240, acquisition time = 4 min. T2-weighted: sequence = CUBE-T2, TR = 2500 ms, TE = 90 ms, echo train length (ETL) = 120, FOV = 256 x 256 mm^2^, matrix size = 320 x 320 pixels, resolution = 0.8 mm x 0.8 mm x 0.8 mm, number of slices = 240, acquisition time = 4 min. The structural MRI data were converted into NIfTI format using the open-source tool *dcm2niix*. Both T1- and T2-weighted images were defaced for all participants using the open-source Python-based defacing utility tool, *pydeface*.

### Sleep Staging

Sleep staging was performed manually in accordance with the American Academy of Sleep Medicine guidelines by a registered polysomnographic technician ^57^. Electrooculography, electromyography and 6 hd-EEG channels that approximate 10:20 locations F3, F4, C3, C4, O1, O2 were used for staging. Hd-EEG data were filtered using MATLAB’s *filtfilt* function with design: 4^th^ order infinite impulse response filter band-passed 1–30 Hz. Electromyography data were filtered using MATLAB’s *filtfilt* function with design: 4^th^ order infinite impulse response filter band-passed 1–100 Hz followed by a notch filter: 4^th^ order infinite impulse response filter stop band 59-61 Hz. CountingSheepPSG (v1.4.5) MATLAB software was then used to perform staging in 30-second epochs and calculate sleep metrics such as Total Sleep Time, Sleep Onset Latency (to Stage 1), Wake After Sleep Onset, and others. Nonparametric statistical tests were used to evaluate for potential changes in these metrics across participant groups (Wilcoxon rank sum) and within participants, across intervention nights (Friedman test).

### hd-EEG Preprocessing

hd-EEG data preprocessing was performed using MATLAB (v2023b) and EEGLAB (v2025.0.0) ^58^. hd-EEG data were band-pass filtered 0.5–40 Hz using EEGLAB’s finite impulse response filter function, *pop_eegfiltnew*. Only NREM epochs staged 2 or 3 were included in spectral power analyses.

### hd-EEG Artifact Rejection

Semi-automated artifact rejection was performed to remove channels and epochs containing artifactual activity (e.g., high-frequency noise, interrupted contact with the scalp) ^59-61^. Specifically, a Fast Fourier Transform (FFT) was used to calculate spectral power in low (0.5–4 Hz) and high (20–30 Hz) frequency ranges for 6-second NREM epochs. For each channel, automatic thresholds were calculated at the 99th percentile and then plotted and visually inspected. If epochs with substantially greater low- or high-frequency power did not exceed the automatic threshold, the threshold was lowered. All epochs exceeding the threshold were then removed.

Channels in which artifacts affected most of the recording were removed. Additional spectral- and topographic-based procedures were used to identify and remove channels with distinctly greater power relative to neighboring channels. On average, 74% (189±23) of the 256-hd EEG channels remained on intervention nights. A final visual review of the hd-EEG data was performed to remove any remaining channels and epochs with artifactual activity.

To increase the signal-to-noise ratio, analyses were restricted to 185 scalp channels (excluding neck and face channels). Any removed channels, including channels that were not gelled, within the scalp 185 channels were interpolated using spherical spline interpolation via the *eeg_interp* function. All channel data was then re-referenced to the average of the final 185 channels. For visualization only, topographies were plotted using EEGLAB’s *topoplot* function with a decreased radius (plotrad = 0.51); all 185 channels were included in analyses.

### hd-EEG Alignment

Timing of TES-TI throughout the night was determined based on hardware trigger signals in agreement with timestamps recorded at the start and end of individual stimulation protocols. Time periods included a baseline PRE stimulation period 3-minutes prior to the start of stimulation, a STIM period including a 15-second ramp up, 3-minute stimulation, and 15-second ramp down, and a POST stimulation period 3-minutes after the ramp down. Analyses comparing STIM and PRE or POST intervals excluded the ramping interval to allow for equal 3-minutes comparisons.

### hd-EEG Spectral Power Analyses

For each hd-EEG channel, power spectral density (PSD) was calculated with Welch’s method using 6-second windows with 90% overlap for a 0.167 Hz step size. PSD values were then averaged over frequencies to calculate band power (e.g., SWA, 0.5–4 Hz). Band power was normalized to account for differences between participants and night-to-night variability within participants ^48^. For each participant and each night, the band power of interest (e.g., SWA) for each channel in PRE, STIM, and POST periods was divided by the average band power (e.g., SWA) during NREM sleep throughout the entire night to obtain a relative change (shown as a percent) from the mean overnight band power ^48^. Canonical frequency bands were defined as follows, SWA: 0.5–4 Hz, theta: 4–8 Hz, alpha: 8–12 Hz, low sigma: 9–12 Hz, high sigma: 12–16 Hz, beta: 16–25 Hz, gamma: 25–40 Hz.

### Statistics

In general, data were evaluated for normality using the Shapiro-Wilk test prior to performing any statistical tests. If data distributions were normal, parametric tests were used; if data distributions were not normal, nonparametric tests were used. Tests were two-tailed unless otherwise specified. Assessment of statistical significance in the context of multiple comparisons was performed using the false discovery rate, particularly the Benjamini-Yekutieli procedure. All results are reported as mean ± standard deviation and all reported p-values are unadjusted unless otherwise specified. For topographic comparisons, a nonparametric cluster-based statistical approach using a suprathreshold cluster analysis to control for multiple comparisons was implemented ^62-64^. For each permutation (N=5000), new datasets were generated with randomly relabeled conditions from the original data. Then, a selected statistical test (e.g., t-test or Spearman correlation) was performed and clusters were determined based on neighboring channels exceeding the critical value for a given two-tailed test distribution, degrees of freedom, and alpha of 0.05. The maximal size of resulting clusters was used to build a cluster size distribution across all permutations. The critical cluster size threshold was defined as the 95^th^ percentile of this distribution. For the empirical comparison, only channels that exceeded the critical value and were located within a cluster larger than the critical cluster size threshold were considered significant ^62,63^. The number of significant clusters, total number of significant electrodes, and mean p-value (p̄) of those significant electrodes are reported; channels with significant responses are shown as white dots in figures.

## Data Availability

The data underlying this article will be shared on reasonable request to the corresponding authors.

## Code Availability

Customized codes used to create the main figures and tables have been made publicly available on GitHub, at https://github.com/eschaeffer-0/Schaeffer_et_al_2025.

## Acknowledgements

We thank Tricia Denman, Emily Peterson, and Kate Riordan for their efforts in participant recruitment, Everett Carstens, Claire Doucet, Paul Ladefoged, Poorang Nori, and Ana Maria Vascan for their efforts in data collection, Cameron Brace and Dillon Scott for their assistance in data processing.

This project is funded by the Defense Advanced Research Projects Agency (DARPA) under cooperative agreement No. HR00112320033 (to G.T., R.J.D.). The content of the information does not necessarily reflect the position or the policy of the Government, and no official endorsement should be inferred.

The TIBS-R investigational devices used in this project were provided by TI Solutions AG, Switzerland, as part of its Early Adopter Program (www.temporalinterference.com).

## Author Contributions

Conceptualization – G.T., R.J.D., S.G.J., R.I.G., C.C., M.B., E.L.S., I.H.; Data Curation – E.L.S., I.H., Z.F., S.Br., B.M., R.S., S.G.J., L.A., M.B.; Formal Analysis – E.L.S., S.Br., I.H., Z.F., L.N.; Funding Acquisition – G.T., R.J.D., S.G.J., R.I.G.; Investigation – E.L.S., I.H., B.M., T.A., G.V., R.S., L.N.; Methodology – G.T., S.G.J., M.B., E.L.S., I.H., Z.F., S.Br., L.A., F.M., A.W., P.A., S.Be., M.C., E.N., N.K.; Project Administration – S.G.J., E.L.S., B.M., R.S., M.B.; Resources – P.A., S.Be., M.C., E.N., N.K.; Software – E.L.S., I.H., Z.F., S.Br., P.A., S.Be., M.C., E.N., N.K.; Visualization – E.L.S., S.Br., I.H., Z.F.; Writing (Original Draft) – E.L.S., C.C., G.T.; Writing (Review & Editing) – all authors.

## References

1. Steriade, M., Timofeev, I. & Grenier, F. Natural waking and sleep states: a view from inside neocortical neurons. Journal of neurophysiology 85, 1969–1985 (2001).

2. Borbély, A.A. A two process model of sleep regulation. Human Neurobiol. 1, 195–204 (1982).

3. Borbely, A.A., Daan, S., Wirz-Justice, A. & Deboer, T. The two-process model of sleep regulation: a reappraisal. J Sleep Res 25, 131–143 (2016).

4. Franken, P. & Dijk, D.J. Sleep and circadian rhythmicity as entangled processes serving homeostasis. Nat Rev Neurosci 25, 43–59 (2024).

5. Gulati, T., Guo, L., Ramanathan, D.S., Bodepudi, A. & Ganguly, K. Neural reactivations during sleep determine network credit assignment. Nature neuroscience 20, 1277–1284 (2017).

6. Liu, J., et al. Slow-wave sleep drives sleep-dependent renormalization of synaptic AMPA receptor levels in the hypothalamus. PLoS Biol 22, e3002768 (2024).

7. Squarcio, F., Tononi, G. & Cirelli, C. Effects of non-rapid eye movement sleep on the cortical synaptic expression of GluA1-containing AMPA receptors. The European journal of neuroscience 60, 3961–3972 (2024).

8. Tononi, G. & Cirelli, C. Sleep and the price of plasticity: from synaptic and cellular homeostasis to memory consolidation and integration. Neuron 81, 12–34 (2014).

9. Landsness, E.C., et al. Sleep-dependent improvement in visuomotor learning: a casual role for slow waves. Sleep 32, 1273–1284 (2009).

10. Bellesi, M., Riedner, B.A., Garcia-Molina, G.N., Cirelli, C. & Tononi, G. Enhancement of sleep slow waves: underlying mechanisms and practical consequences. Front Syst Neurosci 8, 208 (2014).

11. Aeschbach, D., Cutler, A.J. & Ronda, J.M. A role for non-rapid-eye-movement sleep homeostasis in perceptual learning. The Journal of neuroscience: the official journal of the Society for Neuroscience 28, 2766–2772 (2008).

12. Marshall, L., Molle, M., Hallschmid, M. & Born, J. Transcranial direct current stimulation during sleep improves declarative memory. The Journal of neuroscience: the official journal of the Society for Neuroscience 24, 9985–9992 (2004).

13. Wunderlin, M., et al. Acoustic stimulation during sleep predicts long-lasting increases in memory performance and beneficial amyloid response in older adults. Age Ageing 52, afad228 (2023).

14. Feher, K.D., et al. Shaping the slow waves of sleep: A systematic and integrative review of sleep slow wave modulation in humans using non-invasive brain stimulation. Sleep Med Rev 58, 101438 (2021).

15. Luff, C.E. & de Lecea, L. Can Neuromodulation Improve Sleep and Psychiatric Symptoms? Curr Psychiatry Rep 26, 650–658 (2024).

16. Brodt, S., Inostroza, M., Niethard, N. & Born, J. Sleep-A brain-state serving systems memory consolidation. Neuron 111, 1050–1075 (2023).

17. Louviot, S., et al. Transcranial Electrical Stimulation generates electric fields in deep human brain structures. Brain stimulation 15, 1–12 (2022).

18. Grossman, N., et al. Noninvasive Deep Brain Stimulation via Temporally Interfering Electric Fields. Cell 169, 1029–1041 e1016 (2017).

19. Thiele, C., Tamm, C., Ruhnau, P. & Zaehle, T. Perceptibility and Pain Thresholds in Low- and High-Frequency Alternating Current Stimulation: Implications for tACS and tTIS.. Journal of Cognitive Enhancement, 10.1007/s41465-41024-00304-41462 (2024).

20. Vassiliadis, P., et al. Safety, tolerability and blinding efficiency of non-invasive deep transcranial temporal interference stimulation: first experience from more than 250 sessions. J Neural Eng 21(2024).

21. Wang, Y., et al. The safety and efficacy of applying a high-current temporal interference electrical stimulation in humans. Front Hum Neurosci 18, 1484593 (2024).

22. Missey, F., et al. Non-invasive Temporal Interference Stimulation of the Hippocampus Suppresses Epileptic Biomarkers in Patients with Epilepsy: Biophysical Differences between Kilohertz and Amplitude Modulated Stimulation. medRxiv (2025).

23. Beanato, E., et al. Noninvasive modulation of the hippocampal-entorhinal complex during spatial navigation in humans. Sci Adv 10, eado4103 (2024).

24. Thiele, C., Rufener, K.S., Repplinger, S., Zaehle, T. & Ruhnau, P. Transcranial temporal interference stimulation (tTIS) influences event-related alpha activity during mental rotation. Psychophysiology 61, e14651 (2024).

25. Violante, I.R., et al. Non-invasive temporal interference electrical stimulation of the human hippocampus. Nature neuroscience 26, 1994–2004 (2023).

26. Piao, Y., et al. Safety Evaluation of Employing Temporal Interference Transcranial Alternating Current Stimulation in Human Studies. Brain sciences 12, 1194 (2022).

27. Murphy, M.J., et al. Source modeling sleep slow waves. Proceedings of the National Academy of Sciences of the United States of America 106, 1608–1613 (2009).

28. Nir, Y., et al. Regional slow waves and spindles in human sleep. Neuron 70, 153–169 (2011).

29. Robbins, R., et al. A Nationally Representative Survey Assessing Restorative Sleep in US Adults. Front Sleep 1, 935228 (2022).

30. Drake, C.L., et al. Development and evaluation of a measure to assess restorative sleep. J Clin Sleep Med 10, 733–741, 741A-741E (2014).

31. Riedner, B.A., Hulse, B.K., Murphy, M.J., Ferrarelli, F. & Tononi, G. Temporal dynamics of cortical sources underlying spontaneous and peripherally evoked slow waves. Progress in brain research 193, 201–218 (2011).

32. Tononi, G., Riedner, B.A., Hulse, B.K., Ferrarelli, F. & Sarasso, S. Enhancing sleep slow waves with natural stimuli. Medicamundi 54, 73–79 (2010).

33. Ketz, N., Jones, A.P., Bryant, N.B., Clark, V.P. & Pilly, P.K. Closed-Loop Slow-Wave tACS Improves Sleep-Dependent Long-Term Memory Generalization by Modulating Endogenous Oscillations. The Journal of neuroscience: the official journal of the Society for Neuroscience 38, 7314–7326 (2018).

34. Ladenbauer, J., et al. Promoting Sleep Oscillations and Their Functional Coupling by Transcranial Stimulation Enhances Memory Consolidation in Mild Cognitive Impairment. The Journal of neuroscience: the official journal of the Society for Neuroscience 37, 7111–7124 (2017).

35. Siclari, F., et al. Two distinct synchronization processes in the transition to sleep: a high-density electroencephalographic study. Sleep 37, 1621–1637 (2014).

36. Peterchev, A.V. One’s trash is another’s treasure: Subthreshold kilohertz brain modulation as a side effect and as an intervention. Brain stimulation 18, 622–623 (2025).

37. Marzano, C., et al. How we fall asleep: regional and temporal differences in electroencephalographic synchronization at sleep onset. Sleep medicine 14, 1112–1122 (2013).

38. Gorgoni, M., et al. The Regional EEG Pattern of the Sleep Onset Process in Older Adults. Brain sciences 11, 1261 (2021).

39. Borbely, A.A., Baumann, F., Brandeis, D., Strauch, I. & Lehmann, D. Sleep deprivation: effect on sleep stages and EEG power density in man. Electroencephalogr Clin Neurophysiol 51, 483–495 (1981).

40. Maes, J., et al. Sleep misperception, EEG characteristics and autonomic nervous system activity in primary insomnia: a retrospective study on polysomnographic data. Int J Psychophysiol 91, 163–171 (2014).

41. Fernandez, L.M.J. & Luthi, A. Sleep Spindles: Mechanisms and Functions. Physiol Rev 100, 805–868 (2020).

42. Dijk, D.J. EEG slow waves and sleep spindles: windows on the sleeping brain. Behav Brain Res 69, 109–116 (1995).

43. Kesebir, P., Gasiorowska, A., Goldman, R., Hirshberg, M.J. & Davidson, R.J. Emotional Style Questionnaire: A multidimensional measure of healthy emotionality. Psychol Assess 31, 1234–1246 (2019).

44. Martin, M.M. & Rubin, R.B. A New Measure of Cognitive Flexibility. Psych Reports 76, 623–626 (1995).

45. Brahma, T., Guillen, A., Datta, A. & Huang, Y. On the need of individually optimizing temporal interference stimulation of human brains due to inter-individual variability.. bioRxiv. 10.1101/2025.01.13.632831 (2025).

46. Carrier, J., et al. Sleep slow wave changes during the middle years of life. The European journal of neuroscience 33, 758–766 (2011).

47. Mander, B.A., Winer, J.R. & Walker, M.P. Sleep and Human Aging. Neuron 94, 19–36 (2017).

48. Krugliakova, E., et al. Boosting Recovery During Sleep by Means of Auditory Stimulation. Front Neurosci 16, 755958 (2022).

49. Mirzakhalili, E., Barra, B., Capogrosso, M. & Lempka, S.F. Biophysics of Temporal Interference Stimulation. Cell Syst 11, 557–572 (2020).

50. Cassara, A.M., et al. Recommendations for the Safe Application of Temporal Interference Stimulation in the Human Brain Part I: Principles of Electrical Neuromodulation and Adverse Effects. Bioelectromagnetics 46, e22542 (2025).

51. Cassara, A.M., et al. Recommendations for the Safe Application of Temporal Interference Stimulation in the Human Brain Part II: Biophysics, Dosimetry, and Safety Recommendations. Bioelectromagnetics 46, e22536 (2025).

52. Iacono, M.I., et al. MIDA: A Multimodal Imaging-Based Detailed Anatomical Model of the Human Head and Neck. PloS one 10, e0124126 (2015).

53. Windhoff, M., Opitz, A. & Thielscher, A. Electric field calculations in brain stimulation based on finite elements: an optimized processing pipeline for the generation and usage of accurate individual head models. Hum Brain Mapp 34, 923–935 (2013).

54. Saturnino, G.B., Madsen, K.H. & Thielscher, A. Optimizing the electric field strength in multiple targets for multichannel transcranial electric stimulation. J Neural Eng 18, 10.1088 (2021).

55. Cox, R.W. AFNI: software for analysis and visualization of functional magnetic resonance neuroimages. Computers and biomedical research, an international journal 29, 162–173 (1996).

56. Cox, R.W. & Hyde, J.S. Software tools for analysis and visualization of fMRI data. NMR Biomed 10, 171–178 (1997).

57. Berry, R.B., et al. The AASM Manual for the Scoring of Sleep and Associated Events. Rules, terminology and technical specifications. Version 2.5. American Academy of Sleep Medicine, Darien, IL. Available at www.aasmnet.org(2018).

58. Delorme, A. & Makeig, S. EEGLAB: an open source toolbox for analysis of single-trial EEG dynamics including independent component analysis. J Neurosci Methods 134, 9–21. (2004).

59. Jones, S.G., et al. Regional reductions in sleep electroencephalography power in obstructive sleep apnea: a high-density EEG study. Sleep 37, 399–407 (2014).

60. Riedner, B.A., et al. Regional Patterns of Elevated Alpha and High-Frequency Electroencephalographic Activity during Nonrapid Eye Movement Sleep in Chronic Insomnia: A Pilot Study. Sleep 39, 801–812 (2016).

61. Valomon, A., et al. A high-density electroencephalography study reveals abnormal sleep homeostasis in patients with rapid eye movement sleep behavior disorder. Sci Rep 11, 4758 (2021).

62. Fattinger, S., et al. Deep sleep maintains learning efficiency of the human brain. Nature communications 8, 15405 (2017).

63. Huber, R., Ghilardi, M.F., Massimini, M. & Tononi, G. Local sleep and learning. Nature 430, 78–81 (2004).

64. Nichols, T.E. & Holmes, A.P. Nonparametric permutation tests for functional neuroimaging: a primer with examples. Hum Brain Mapp 15, 1–25 (2002).

