## supplementary information for "Enhancement of Sleep Slow Wave Activity using Transcranial Electrical Stimulation with Temporal Interference"

**Supplementary Information (1 figure, 3 tables)**

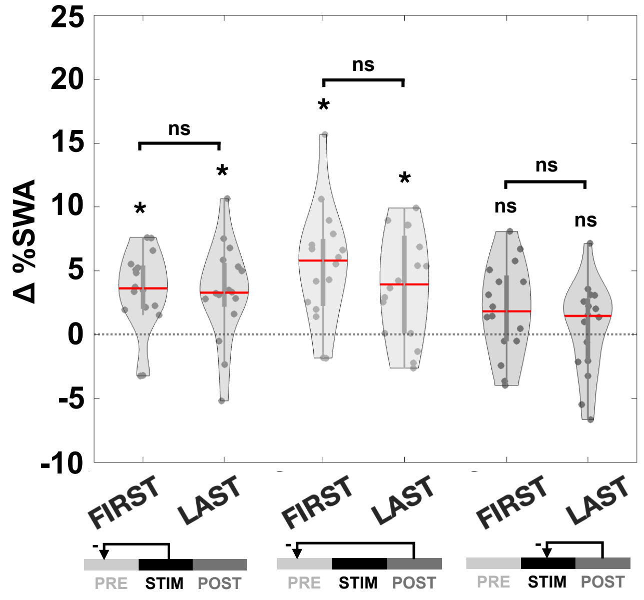


***Supplementary Figure 1. Consistent TES$^{15kHz}$-TI$^{1Hz}$ effects on first and last intervention nights***. *For participants with usable data during TES$^{15kHz}$-TI$^{1Hz}$ stimulation on both first and last nights (N=16), across all channels, there was a main effect with a significant difference in the average normalized SWA (%SWA) across PRE, STIM, and POST periods (df=2, F=25.20, p=3.79x10$^{-7}$), but there was no main effect across nights (df=1, F=0.31, p=0.584) and there was no interaction effect between time period and night (df=2, F=0.82, p=0.448) when evaluated with a two-way repeated measures ANOVA. (Left) %SWA during stimulation compared to before stimulation (STIM-PRE) showed similar increases on the first (N=16, t=4.24, p=7.09x10$^{-4}$, d=1.06) and last (t=3.50, p=3.24x10-3, d=0.87) intervention night, with no significant difference between nights (t=0.04, p=0.517, d=0.01). (Middle) Increases in %SWA after stimulation compared to before stimulation (POST-PRE) were also similar on the first (N=16, t=4.79, p=2.40x10$^{-4}$, d=1.20) and last (t=3.67, p=2.26x10$^{-3}$, d=0.92) intervention night, with no significant difference between nights (t=1.02, p=0.841, d=0.25). (Right) %SWA did not differ after stimulation compared to during stimulation (POST-STIM, either on the first (N=16, t=1.93, p=0.046, adjusted p=0.136, d=0.47) or the last (t=0.52, p=0.612, d=0.13) intervention night, with no significant difference between nights (t=1.18, p=0.876, d=0.29). Two-tailed paired samples t-tests were used. Median shown as solid red line.*

***Supplementary Table 1. Sleep characteristics across nights in TES$^{15kHz}$.*** *Values are Mean ± standard deviation.*

| | Baseline Night | First Night | Last Night | Test Statistic ($X^2$), p-value |
|---|---|---|---|---|
| **Total Sleep-Wake Period (hr)** | 7.74 ± 1.16 | 7.62 ± 0.52 | 7.49 ± 0.23 | 0.40, 0.819 |
| **Total Sleep Time (hr)** | 6.49 ± 1.48 | 6.45 ± 1.40 | 6.76 ± 0.82 | 1.20, 0.549 |
| **Sleep Onset Latency (min)** | 29.50 ± 13.52 | 18.30 ± 13.67 | 11.90 ± 5.53 | 8.40, 0.015$^{\dagger}$ |
| **Sleep Efficiency (%)** | 83.46 ± 12.35 | 84.38 ± 15.96 | 90.13 ± 8.61 | 3.60, 0.165 |
| **Number of Awakenings** | 11 ± 6 | 14 ± 5 | 18 ± 7 | 2.80, 0.247 |
| **Wake After Sleep Onset (min)** | 45.10 ± 43.64 | 51.50 ± 58.02 | 31.30 ± 31.57 | 0.40, 0.819 |
| **Stage N1 (%)** | 2.45 ± 2.16 | 2.90 ± 1.80 | 3.97 ± 4.49 | 0.40, 0.819 |
| **Stage N2 (%)** | 61.38 ± 4.47 | 64.87 ± 7.52 | 66.85 ± 4.46 | 1.60, 0.449 |
| **Stage N3 (%)** | 16.72 ± 4.00 | 10.74 ± 4.11 | 9.81 ± 6.49 | 5.20, 0.074 |
| **Stage REM (%)** | 19.45 ± 4.19 | 21.49 ± 7.06 | 19.38 ± 2.81 | 1.20, 0.549 |

***Supplementary Table 2. Statistics for TES$^{15kHz}$-TI$^{1Hz}$ effects on other frequency bands.*** *For each canonical frequency band, average power spectral density (PSD) values were compared during (STIM-PRE) and after (POST-PRE) stimulation as compared to before stimulation for TES$^{15kHz}$-TI$^{1Hz}$ participants (N=21). Paired samples t-tests were used with the Benjamini-Yekutieli procedure to correct for multiple comparisons. T-statistics, unadjusted p-values, and adjusted p-values reported. As compared to before stimulation, during (STIM-PRE), in addition to SWA (Figure 2), theta power increased, and beta power decreased. There was also a decrease in high sigma power, but it did not pass correction for multiple comparisons. Similarly, after stimulation (POST-PRE), in addition to SWA (Figure 2), theta power increased, and high sigma power decreased.*

| | STIM-PRE | | | POST-PRE | | |
|---|---|---|---|---|---|---|
| | **T-Statistic** | **P-Value** | **Adjusted P-Value** | **T-Statistic** | **P-Value** | **Adjusted P-Value** |
| **Theta (4–8 Hz)** | 4.29 | $3.58 \times 10^{-4}$ | $8.12 \times 10^{-3}$ | 3.90 | $8.92 \times 10^{-4}$ | $8.12 \times 10^{-3}$ |
| **Alpha (8–12 Hz)** | 1.75 | 0.096 | 0.438 | 1.34 | 0.194 | 0.803 |
| **Sigma (Low, 9–12 Hz)** | 0.68 | 0.502 | 1.632 | 0.80 | 0.433 | 1.516 |
| **Sigma (High, 12–16 Hz)** | -2.75 | 0.012 | 0.080 | -3.95 | $7.95 \times 10^{-4}$ | $8.12 \times 10^{-3}$ |
| **Beta (16–25 Hz)** | -3.29 | $3.68 \times 10^{-3}$ | 0.028 | -1.96 | 0.064 | 0.324 |
| **Gamma (25–40 Hz)** | -2.06 | 0.053 | 0.301 | -1.23 | 0.233 | 0.883 |

***Supplementary Table 3. Summary of Figure Statistics.*** *95% Confidence Intervals (CI) shown. All tests were two-sided. Adjustment for multiple comparisons was performed using the Benjamini-Yekutieli procedure.*

| Analysis | Figure | Number of Participants | Median, [Lower CI, Upper CI] | Statistical Test | Degrees of Freedom | Test Statistic | Unadjusted P-Value | Adjusted P-Value | Effect Size (Cohen's d) |
|---|---|---|---|---|---|---|---|---|---|
| PRE, STIM, POST (%SWA) | Fig 2A | $TES^{15kHz}$-$TI^{1Hz}$ = 21 | | One-Way Repeated Measures ANOVA | 2 | F = 30.97 | **7.48x$10^{-9}$** | | |
| STIM-PRE (Δ%SWA) | Fig 2A | $TES^{15kHz}$-$TI^{1Hz}$ = 21 | 3.48, [2.68, 4.75] | Paired T-Test | 20 | T = 6.64 | **1.81x$10^{-6}$** | **4.99x$10^{-6}$** | 1.45 |
| POST-PRE (Δ%SWA) | Fig 2A | $TES^{15kHz}$-$TI^{1Hz}$ = 21 | 4.61, [3.64, 6.13] | Paired T-Test | 20 | T = 7.47 | **3.34x$10^{-7}$** | **1.83x$10^{-6}$** | 1.63 |
| POST-STIM (Δ%SWA) | Fig 2A | $TES^{15kHz}$-$TI^{1Hz}$ = 21 | 1.42, [-1.19, 1.57] | Paired T-Test | 20 | T = 0.63 | 0.536 | 0.982 | 0.14 |
| STIM (%SWA) | Fig 3B | $TES^{15kHz}$-$TI^{1Hz}$ = 21<br>$TES^{15kHz}$ = 7 | $TES^{15kHz}$-$TI^{1Hz}$ = 104.63, [103.32, 106.19]<br>$TES^{15kHz}$ = 98.85, [97.48, 104.73] | Independent T-Test | 26 | T = 2.48 | **0.020** | | 1.08 |
| STIM EARLY v. LATE (Δ%SWA) | Fig 3D | $TES^{15kHz}$-$TI^{1Hz}$ = 21 | 3.76, [0.69, 4.16] | Paired T-Test | 20 | T = 3.25 | **4.02x$10^{-3}$** | | 0.71 |
| STIM EARLY v. LATE (Δ%SWA) | Fig 3D | $TES^{15kHz}$ = 7 | 0.75, [0.17, 5.18] | Paired T-Test | 6 | T = 1.71 | 0.138 | | 0.65 |
| STIM-PRE (Theta ΔPSD) | Fig 4A | $TES^{15kHz}$-$TI^{1Hz}$ = 21 | 0.15, [0.11, 0.28] | Paired T-Test | 20 | T = 4.29 | **3.58x$10^{-4}$** | **8.12x$10^{-3}$** | 0.94 |
| STIM-PRE (Beta ΔPSD) | Fig 4A | $TES^{15kHz}$-$TI^{1Hz}$ = 21 | 0.17, [0.12, 0.36] | Paired T-Test | 20 | T = -3.29 | **3.68x$10^{-3}$** | **0.028** | -0.72 |
| POST-PRE (Theta ΔPSD) | Fig 4C | $TES^{15kHz}$-$TI^{1Hz}$ = 21 | -4.39x$10^{-3}$, [-1.11x$10^{-2}$, -3.56x$10^{-3}$] | Paired T-Test | 20 | T = 3.90 | **8.92x$10^{-4}$** | **8.12x$10^{-3}$** | 0.85 |
| POST-PRE (Sigma ΔPSD) | Fig 4C | $TES^{15kHz}$-$TI^{1Hz}$ = 21 | -3.12x$10^{-2}$, [-7.06x$10^{-2}$, -2.63x$10^{-2}$] | Paired T-Test | 20 | T = -3.95 | **7.95x$10^{-4}$** | **8.12x$10^{-3}$** | -0.86 |
| ΔREST-Q v. Δ%SWA | Fig 5A | $TES^{15kHz}$-$TI^{1Hz}$ = 16 | | Spearman's Rho | 14 | Rs = 0.56 | **0.023** | | |
| ΔREST-Q v. Δ%Low Sigma | Fig 5B | $TES^{15kHz}$-$TI^{1Hz}$ = 16 | | Spearman's Rho | 14 | Rs = 0.15 | 0.573 | | |
| ΔREST-Q v. Δ%High Sigma | Fig 5C | $TES^{15kHz}$-$TI^{1Hz}$ = 16 | | Spearman's Rho | 14 | Rs = -0.23 | 0.393 | | |
| First v. Last: PRE, STIM, POST (%SWA) | Suppl Fig 1 | $TES^{15kHz}$-$TI^{1Hz}$ = 16 | | Two-Way Repeated Measures ANOVA | 2 | F = 25.20 | **3.79x$10^{-7}$** | | |
| First v. Last: First or Last Night | Suppl Fig 1 | $TES^{15kHz}$-$TI^{1Hz}$ = 16 | | Two-Way Repeated Measures ANOVA | 1 | F = 0.31 | 0.584 | | |
| First v. Last: Interaction | Suppl Fig 1 | $TES^{15kHz}$-$TI^{1Hz}$ = 16 | | Two-Way Repeated Measures ANOVA | 2 | F = 0.82 | 0.448 | | |
| First STIM-PRE (Δ%SWA) | Suppl Fig 1 | $TES^{15kHz}$-$TI^{1Hz}$ = 16 | 3.62, [1.68, 4.72] | Paired T-Test | 15 | T = 4.24 | **7.09x$10^{-4}$** | **5.21x$10^{-3}$** | 1.06 |
| Last STIM-PRE (Δ%SWA) | Suppl Fig 1 | $TES^{15kHz}$-$TI^{1Hz}$ = 16 | 3.28, [1.26, 4.85] | Paired T-Test | 15 | T= 3.50 | **3.24x$10^{-3}$** | **0.012** | 0.87 |
| First v. Last STIM-PRE (Δ%SWA) | Suppl Fig 1 | $TES^{15kHz}$-$TI^{1Hz}$ = 16 | -0.44, [-2.33, 2.49] | Paired T-Test | 15 | T = 0.04 | 0.517 | | 0.01 |
| First POST-PRE (Δ%SWA) | Suppl Fig 1 | $TES^{15kHz}$-$TI^{1Hz}$ = 16 | 5.80, [3.54, 7.92] | Paired T-Test | 15 | T = 4.79 | **2.40x$10^{-4}$** | **3.53x$10^{-3}$** | 1.20 |
| Last POST-PRE (Δ%SWA) | Suppl Fig 1 | $TES^{15kHz}$-$TI^{1Hz}$ = 16 | 3.93, [1.70, 5.80] | Paired T-Test | 15 | T = 3.67 | **2.26x$10^{-3}$** | **0.011** | 0.92 |
| First v. Last POST-PRE(Δ%SWA) | Suppl Fig 1 | $TES^{15kHz}$-$TI^{1Hz}$ = 16 | 1.85, [-1.68, 4.30] | Paired T-Test | 15 | T = 1.02 | 0.841 | | 0.25 |
| First POST-STIM (Δ%SWA) | Suppl Fig 1 | $TES^{15kHz}$-$TI^{1Hz}$ = 16 | 1.82, [0.34, 3.63] | Paired T-Test | 15 | T = 1.93 | **0.046** | 0.136 | 0.47 |
| Last POST-STIM (Δ%SWA) | Suppl Fig 1 | $TES^{15kHz}$-$TI^{1Hz}$ = 16 | 1.46, [-1.39, 1.98] | Paired T-Test | 15 | T = 0.52 | 0.612 | 1.500 | 0.13 |
| First v. Last POST-STIM(Δ%SWA) | Suppl Fig 1 | $TES^{15kHz}$-$TI^{1Hz}$ = 16 | 1.24, [-0.85, 4.11] | Paired T-Test | 15 | T = 1.18 | 0.876 | | 0.29 |
